# Image-based models of T-cell distribution identify a clinically meaningful response to a dendritic cell vaccine in patients with glioblastoma

**DOI:** 10.1101/2023.07.13.23292619

**Authors:** Kamila M. Bond, Lee Curtin, Andrea Hawkins-Daarud, Javier C. Urcuyo, Gustavo De Leon, Kyle W. Singleton, Ariana E. Afshari, Lisa E. Paulson, Christopher P. Sereduk, Kris A. Smith, Peter Nakaji, Leslie C. Baxter, Devi Prasad Patra, Michael P. Gustafson, Allan B. Dietz, Richard S. Zimmerman, Bernard R. Bendok, Nhan L. Tran, Leland S. Hu, Ian F. Parney, Joshua B. Rubin, Kristin R. Swanson

## Abstract

**BACKGROUND:** Glioblastoma is an extraordinarily heterogeneous tumor, yet the current treatment paradigm is a “one size fits all” approach. Hundreds of glioblastoma clinical trials have been deemed failures because they did not extend median survival, but these cohorts are comprised of patients with diverse tumors. Current methods of assessing treatment efficacy fail to fully account for this heterogeneity.

**METHODS:** Using an image-based modeling approach, we predicted T-cell abundance from serial MRIs of patients enrolled in the dendritic cell (DC) vaccine clinical trial. T-cell predictions were quantified in both the contrast-enhancing and non-enhancing regions of the imageable tumor, and changes over time were assessed.

**RESULTS:** A subset of patients in a DC vaccine clinical trial, who had previously gone undetected, were identified as treatment responsive and benefited from prolonged survival. A mere two months after initial vaccine administration, responsive patients had a decrease in model-predicted T-cells within the contrast-enhancing region, with a simultaneous increase in the T2/FLAIR region.

**CONCLUSIONS:** In a field that has yet to see breakthrough therapies, these results highlight the value of machine learning in enhancing clinical trial assessment, improving our ability to prospectively prognosticate patient outcomes, and advancing the pursuit towards individualized medicine.

## Introduction

Glioblastoma (GBM) is the most common primary brain tumor in adults and has a dismal median survival time of about 16 months.^1–3^ GBMs have innate and adaptive resistance mechanisms that enable them to persist despite aggressive multimodal treatments including surgery, chemotherapy, and irradiation. For example, immunotherapy is showing great promise in multiple cancers but not GBM.^4^ Among the reasons is the well-described immunosuppressive microenvironment involving myeloid-derived suppressor cells and T-cell exhaustion.^5^ While therapeutics are available to target immune suppression, the blood-brain barrier (BBB) further limits the entry of immune cells, immune mediators, and many immunotherapeutic drugs into the central nervous system.^6^ Obstacles such as these critically hinder patients’ ability to mount a robust anti-tumoral immune response.

One strategy to circumvent GBM’s relative immune privilege is to exploit dendritic cells and artificially stimulate the systemic anti-tumor response.^7–9^ These potent antigen-presenting cells can be incubated with allogeneic tumor lysate *in vitro* to sensitize them to GBM antigens. When introduced to the patient in the form of a vaccine, the mature dendritic cells present processed tumor antigen to naïve T-cells and provide signals to promote T-cell differentiation and activation. Activated T-cells, unlike their resting counterparts, are able to cross the BBB that surrounds post-capillary venules which allows them to extravasate into the tumor and surrounding parenchyma.^10^ The goal of dendritic cell vaccine therapy is to produce a coordinated, anti-tumoral, adaptive immune response.

A challenge in the assessment of novel therapies like the dendritic cell vaccine is the ability to quantify treatment responsiveness in light of GBM’s profound heterogeneity. GBM is comprised of a spectrum of genotypes and phenotypes that can be seen both within and between patients.^11,12^ Most clinical trials for GBM are considered to have failed because patients’ survival, on average, remains poor.^13,14^ However, some subsets of patients likely benefit from these experimental therapies but remain undetected when aggregated with dissimilar patients for whom the therapy was not advantageous. Here we report an image-based machine learning model (radiomics) approach to quantify changes in intratumoral T-cell abundance over the course of dendritic cell vaccine treatment. We identify patients who had an early, measurable immune response to the vaccine and benefited from prolonged survival.

## Methods

### Image-localized biopsies and RNA sequencing

All data were collected in accordance with a protocol approved by the Mayo Clinic Institutional Review Board. Patients with clinically-suspected GBM were recruited and were enrolled in the study after obtaining informed consent. Forty-three patients (26M, 17F) underwent surgical resection for primary or recurrent high-grade glioma. A total of 147 biopsies (median, 3 per patient) were harvested under intraoperative stereotactic guidance, and the MRI coordinates of each sample were recorded. A summary of the patient population and biopsy characteristics is provided in **Supplementary Table 1**. Tissue was flash-frozen in liquid nitrogen. RNA sequencing libraries were prepared using the Illumina TruSeq v2 RNAseq kit. Paired-end reads were obtained with the Illumina HiSeq 4000 sequencer, and FASTQ files were aligned to the Gencode human GRCh38.p37 reference genome with STAR V2.7.1a^35^ using a docker RNA-seq pipeline.^36^ Counts were tabulated using htseq-count^37^, corrected for batch effects using ComBat-Seq^38^, and normalized in the unit of transcripts per million (TPM).

### Cellular deconvolution and T-cell estimation

CIBERSORTx^21^ was used to infer the absolute abundance of T-cells in each sample from the RNA expression matrix (**Supplementary Table 2**). The original CIBERSORT leukocyte gene signature (LM22), which distinguishes between 22 human hematopoietic cell types and has been robustly benchmarked, was used for cellular deconvolution.^34^ The LM22 signature classifies T-cells into seven subtypes and activation states. The sum of these seven subtypes was used in downstream analyses to represent total T-cell abundance. CIBERSORTx was performed in absolute mode with B mode batch correction (to account for the application of RNAseq data to a signature that was derived with microarray), quantile normalization disabled, and in 500 permutations. A ceiling was applied such that T-cell outliers (>97.5^th^ percentile) were reassigned to equal the 97.5^th^ percentile.

### Image-localized biopsy feature extraction

All patients had pre-treatment MR imaging that included T1-weighted with gadolinium contrast (T1Gd) and T2-weighted (T2) sequences. Preprocessing of images included co-registration using SimpleElastix^39^, inhomogeneity correction using the N4 algorithim^40^, and whole-brain normalization using brain masks extracted using MONSTR^41^. Spherical ROIs centered at the intraoperatively-recorded biopsy coordinate (diameter, 5mm) were generated using the neuroim package (v0.0.6) in R. A total of 792 first-order and filtered (exponential, logarithm, square, square root, and wavelet) imaging features were extracted from each sequence (T1Gd and T2) within the ROI using PyRadiomics^22^ (v3.0.1, Python 3.7.4).

### Machine learning model generation

All machine learning analyses were performed in R (v4.0.1)^42^ using the package Caret (v6.0-86).^43^ One model was generated using all samples, and two sex-specific models were generated using male and female samples separately. Imaging features that were not correlated with T-cell predictions (p>0.1) or were highly correlated with one another (R>0.9) were excluded. Final models were constructed using 46 and 42 imaging features in the male and female models, respectively (**Supplementary Table 3**). A linear support vector machine regression model (svmLinear) was used to predict T-cell abundance from imaging features. Samples were randomly split 70/30% into training and testing sets. Five-fold cross-validation was repeated ten times on the training set. Models were applied to the unseen test data, and the highest-performing model was selected based on RMSE values and actual vs. predicted correlations in the training and testing scenarios. Using Caret’s built-in variable importance function, the influence of each imaging feature on the model’s prediction was quantified. An evaluation of the image-based model characteristics and the reporting thereof is provided in **Supplementary Table 5**.

### Dendritic cell vaccine trial

Twenty adult patients enrolled in the dendritic cell vaccine clinical trial (NCT01957956).^23^ All subjects had newly diagnosed grade IV astrocytoma and had completed standard treatment of tumor resection or biopsy followed by radiation and temozolomide. A summary of patient demographics is provided in **Supplementary Table 4**. Dendritic cell vaccines were prepared as previously described.^9^ Mononuclear cells were collected by apheresis and CD14^+^ monocytes were isolated by flow cytometry. Autologous dendritic cells were sensitized through incubation with allogeneic tumor lysate from two human-derived GBM cell lines. Patients received intradermal injections of the dendritic cell vaccine for up to twelve cycles. Patients underwent standard MR imaging on the day of the first vaccine dose (“pre-treatment”) and two months thereafter (“post-treatment”).

### Model application to dendritic cell vaccine trial images

Trained individuals used an in-house tumor segmentation software to delineate ROIs on pre- and post-treatment MRIs from patients enrolled in the dendritic cell vaccine clinical trial. Each patient had both a T1Gd and T2/FLAIR ROI for each time point. PyRadiomics^22^ (v3.0.1, Python 3.7.4) was used to calculate voxelwise imaging features within the ROIs with a 5x5 window. The machine learning models used these voxelwise features to predict T-cells across the patient’s tumor. The sum of T-cell predictions across an entire ROI was used to quantify total T-cell abundance.

### Survival analyses

Overall survival was defined as the number of months between the first vaccine dose and the last follow-up or death. Patients were categorized into four T-cell response quadrants (combinations of “increasing” and “decreasing” T-cell groups within the T1Gd and T2/FLAIR regions, calculated by the difference between their post- and pre-treatment T-cell abundance). Cases with a net change of < 2.5% were considered to be T-cell stable, and these cases were combined with those in the “decreasing” group for all subsequent analyses. Survival between two groups was compared using unadjusted and age-adjusted Kaplan-Meier analyses using the package “survival”.^44^ Multivariable Cox proportional hazards analysis was performed to determine the influence of T-cell “quadrant” on patient survival with patient age and sex, extent of tumor resection, IDH mutation status, and MGMT methylation status as covariates.

## Results

### Sex-specific models outperform a sex-agnostic model in the prediction of T-cell abundance from imaging

Medical imaging is the foundation of disease monitoring for patients with brain tumors. Radiomics models can be trained to use complex features from these images to detect and predict biological characteristics. We take a particular interest in the imaging representations associated with T-cells, as their dynamics are hypothesized to change as a result of immunotherapies like dendritic cell vaccines.^15^ In acknowledgment of the many described sex differences that exist in GBM biology, the immune system, and imaging^16–20^, we constructed sex-specific radiomics models to predict T-cell abundance from imaging features (**Figure 1A**).

**Figure 1.**
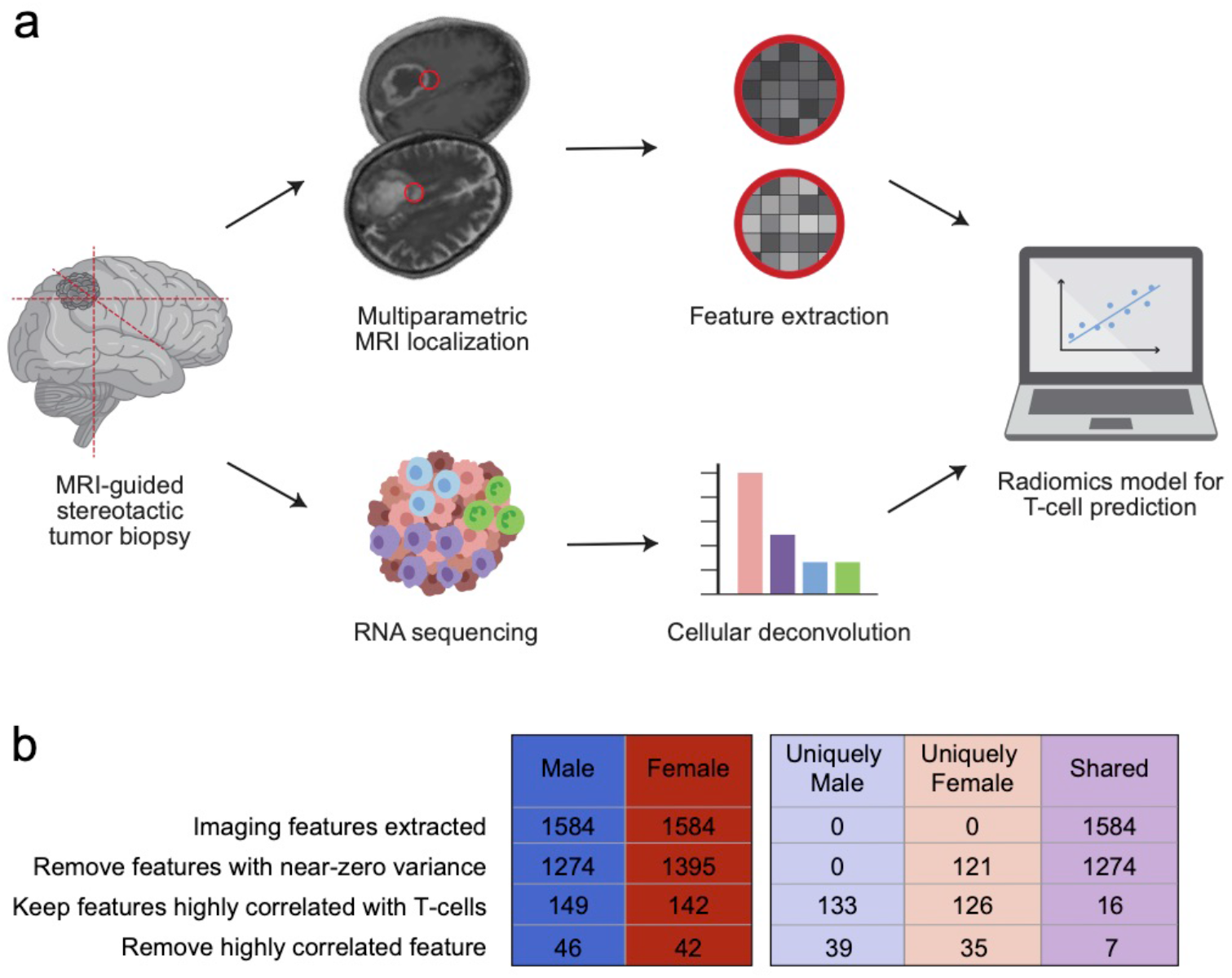
Image-localized biopsies allow for the construction of sex-specific radiomics models that predict T-cells. (a) Under stereotactic guidance, surgeons record the location of tumor biopsies. Imaging features at the biopsy coordinate (red circle) are calculated across multiple MRI sequences. In parallel, the biopsy is sent for RNA sequencing, and deconvolution estimates the abundance of cell types within the sample. Image-based models are built to predict T-cell abundance from imaging features. (b) 1584 imaging features were extracted from each biopsy coordinate. After three feature reduction steps, 46 and 42 imaging features were used to construct male- and female-specific radiomics models, respectively. 39 imaging features were unique to the male model, 35 unique to the female model, and 7 used in both.

Image-localized biopsies (n=147; 81M, 66F) were collected from 43 patients (26M, 17F) with primary or recurrent high-grade glioma (**Supplementary Table 1**). Tissue samples underwent RNA sequencing followed by cellular deconvolution^21^ for the estimation of T-cell abundance (**Supplemental Table 2**). A total of 1584 first-order and filtered imaging features^22^ from T1-weighted with gadolinium (T1Gd) and T2-weighted MRIs were extracted from the biopsy coordinate. After numerous feature reduction steps, male and female-specific models were trained using 46 and 42 imaging features, respectively (**Figure 1B; Supplementary Table 3**).

Samples’ absolute T-cell abundances, as determined by deconvolution, were scaled from 0 to 1 (zero being no T-cells and one being the maximum number of T-cells seen across our entire sample cohort). Support vector machine linear regression models were constructed to predict T-cell abundance from imaging features. Each model was trained using 70% of the cohort and assessed with the remaining unseen test data (**Figure 2A**). The male model performed well on unseen test data, with a Pearson correlation coefficient (R) of 0.67 and a root mean square error (RMSE) of 0.218. Similarly, the female model had R of 0.69 and a RMSE of 0.164 on unseen data.

**Figure 2.**
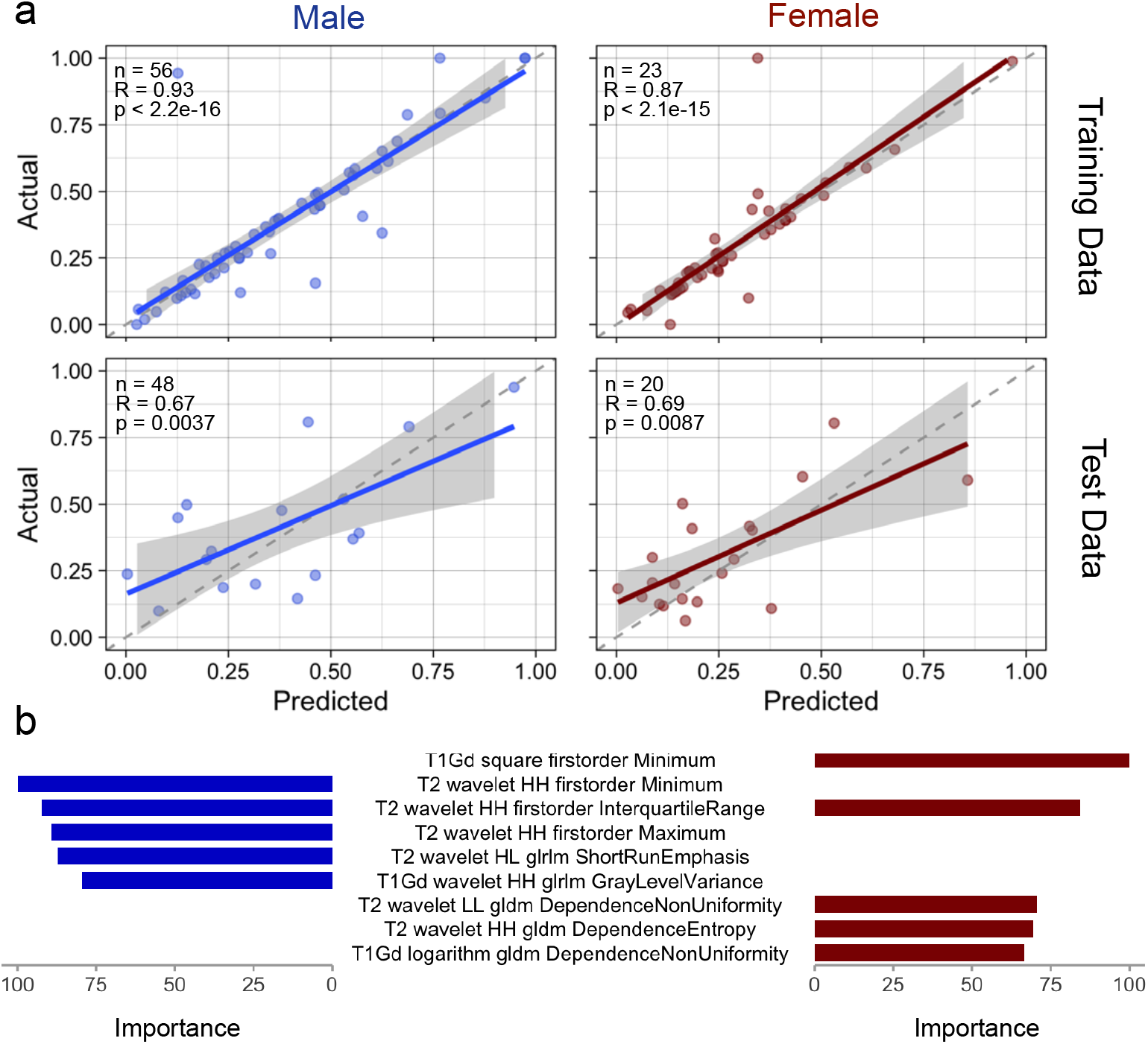
Sex-specific radiomics models use different imaging features in the prediction of T-cells. (a) Male samples (n=81) and female samples (n=66) were independently used to build T-cell prediction models. Each was trained on 70% of the data using a linear support vector machine regression model. Model performance was tested on the remaining 30% of the data. (b) Variable importance plots show the top five imaging features used by the male- and female-specific models.

The sex-specific models outperformed a separate, sex-agnostic model that was trained with all data (Supplementary Figure 1). The RMSE of the combined model performance on unseen test data was 0.370 with an R of 0.65. This indicates that the imaging representations associated with T-cells could be different between males and females. This conclusion was supported by calculations of imaging feature importance, which revealed that the most important variable in the male model was T2 minimum, whereas T1Gd minimum was the most important variable in the female model (**Figure 2B**). These findings support the notion that bioimaging relationships in males and females cannot be assumed to be the same, and sex-specific approaches should be employed when possible.

### The dendritic cell vaccine, on average, provides no clear survival benefit in patients with primary GBM

Our image-based models were applied to an early phase 1 dendritic cell vaccine clinical trial (NCT01957956) as a test use case to evaluate predicted T-cell dynamics over the course of immunotherapy (**Figure 3A**).^23^ Twenty adults with glioblastoma were enrolled. The average age was 61±11 years and 12 (60%) patients were men. The cohort had an overrepresentation of unfavorable prognostic factors including 95% IDH wild type, 70% MGMT unmethylated, and 50% subtotal resection or biopsy (**Supplementary Table 4**). Patients underwent standard-of-care surgical intervention, chemotherapy, and radiation (4 months) before their first vaccine dose. From the time of the first vaccine administration, the median overall survival time was 13 months. While patients’ survival was modestly prolonged compared to historical controls, particularly in light of the cohort’s prognostically unfavorable characteristics, the overall results were underwhelming.^2^

**Figure 3.**
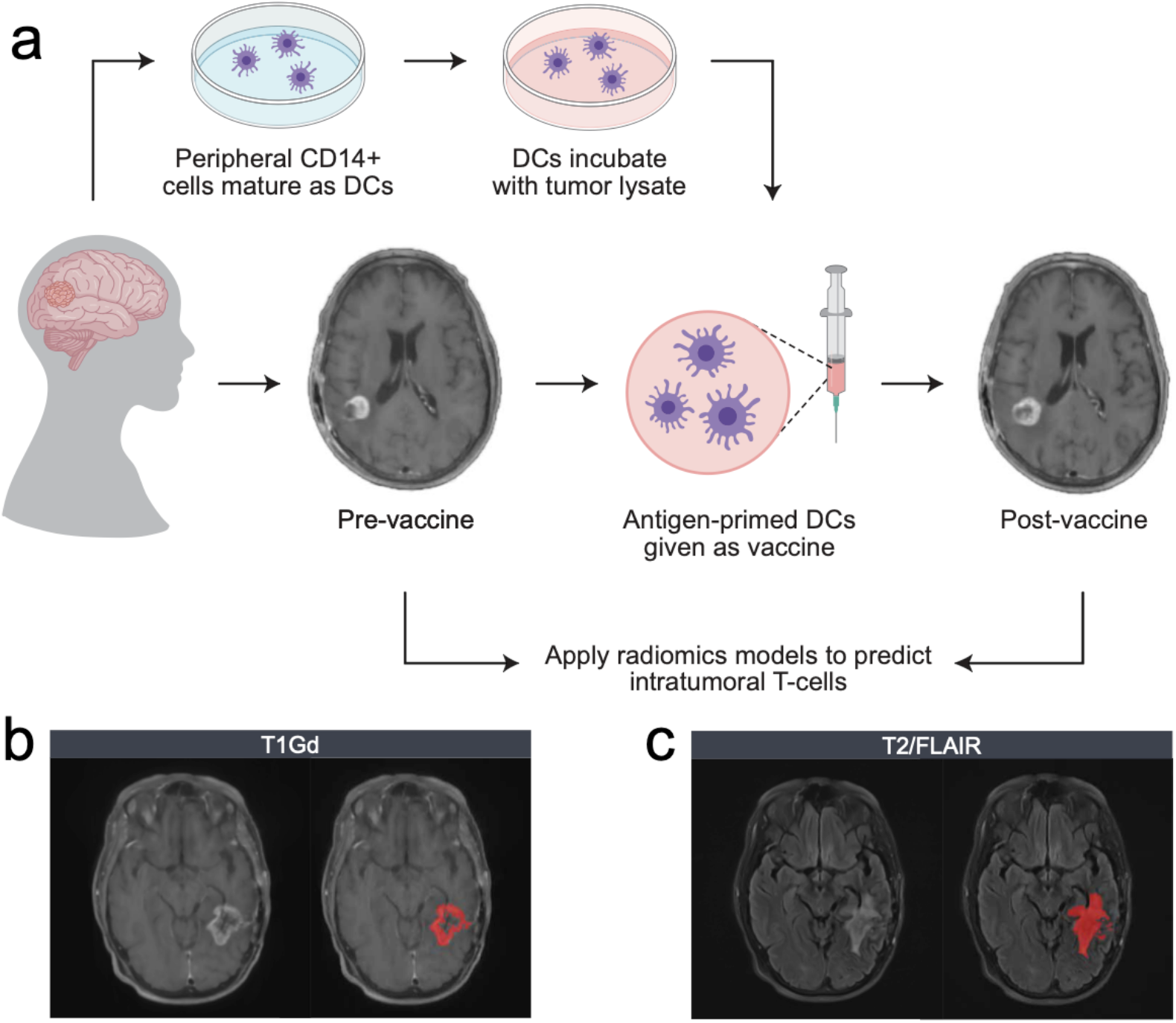
Study design of the dendritic cell (DC) vaccine clinical trial for the treatment of newly diagnosed glioblastoma. (a) Patients with primary glioblastoma underwent standard treatment of surgery, chemotherapy, and radiation. CD14^+^ cells were isolated from peripheral blood, matured into DCs, and incubated with tumor lysate. Sensitized DCs were administered to patients in the form of an adjuvant vaccine. At the pre-vaccine (t=0) and post-vaccine (t=2 months) imaging time points, (b) T1Gd and (c) T2/FLAIR images were obtained and tumors were manually segmented for the voxelwise prediction of intratumoral T-cells.

### Image-based models elucidate spatial patterns of T-cell dynamics related to survival in dendritic cell vaccine patients

As a part of standard disease monitoring, trial patients underwent T1Gd and T2/FLAIR MR imaging (**Figure 3B**) on the date of their first vaccine (pre-treatment) and two months thereafter (post-treatment). To quantify imaging-predicted T-cell dynamics over the course of therapy, we applied our models to these routine images to predict T-cell abundance at each intratumoral voxel within T1Gd and T2/FLAIR regions (**Figure 4A**). Total abundance was calculated as the sum of T-cell predictions across the entire region of interest (ROI) at each time point (**Figure 4B**). The change in total T-cell abundance between imaging time points (post-pre) classified patients into “increasing” and “decreasing” groups. An absolute pre-to-post treatment change of < 2.5% was considered stable, and these cases were combined with those in the “decreasing” category for further analysis. Patients were separated into four categories (**Figure 5A**; combinations of increasing and decreasing/stable within both T1Gd and FLAIR ROIs).

**Figure 4.**
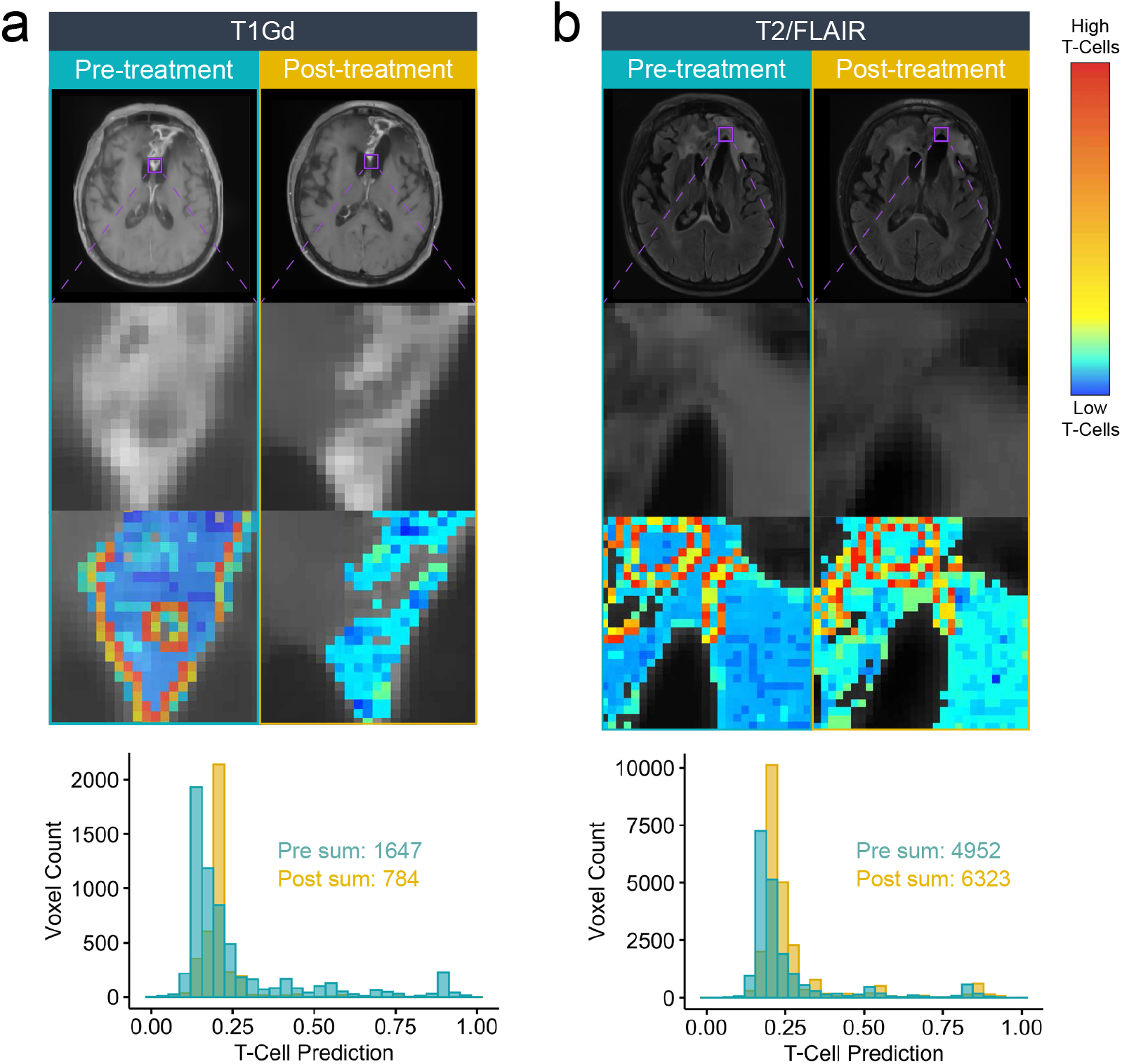
Sex-specific radiomics models make voxelwise T-cell predictions across the imageable tumor. (a) T1Gd and (b) T2/FLAIR MRIs, representative insets (purple squares), and voxelwise T-cell prediction heatmaps are illustrated for the same patient across pre- and post-treatment settings. Histograms represent all T-cell predictions across the entire ROI under each condition. Total T-cell abundance is the sum of the predictions. This patient had a simultaneous decrease in T-cell abundance in the T1Gd region and an increase in T-cell abundance in the T2/FLAIR region.

**Figure 5.**
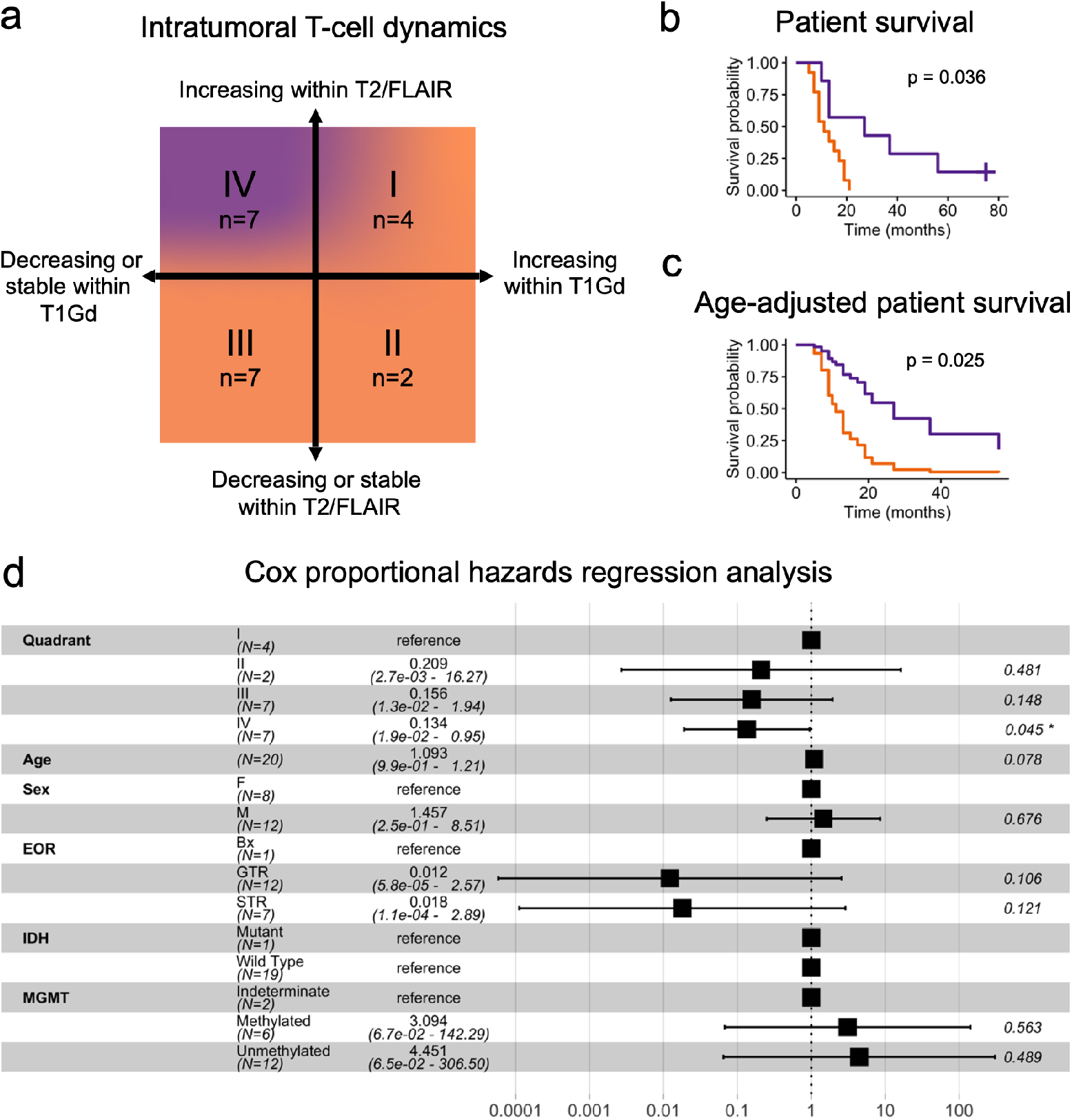
Spatiotemporal T-cell dynamics in response to the dendritic cell vaccine stratify patient survival. (a) Dendritic cell vaccine clinical trial patients (n=20) were assigned into four categories based upon the change in total T-cell predictions within their T1Gd and T2/FLAIR ROIs over the course of treatment. (b) Unadjusted and (c) age-adjusted Kaplan-Meier survival analyses of patients with decreasing T-cells in the T1Gd region with simultaneously increasing T-cells in the T2/FLAIR region compared to all others. (d) Forest plot for Cox proportional hazard model adjusting for patient age, sex, the extent of tumor resection (EOR), IDH mutation status, and MGMT methylation status.

Survival analyses were performed to determine the clinical relevance of the four groups. Kaplan-Meier analysis revealed that patients whose T-cells decreased in the T1Gd ROI while simultaneously increasing in the T2/FLAIR ROI (n=7) survived longer (median, 27 months) than all other patients (p=0.0036; **Figure 5B**). The significance remained in an age-adjusted Kaplan-Meier comparison (p=0.0245; **Figure 5C**), as well as a Cox proportional hazard model that adjusted for patient age and sex, extent of tumor resection, IDH mutation status, and MGMT methylation status (p=0.045; **Figure 5D**). These results suggest that spatiotemporal T-cell modeling within the tumor identified a subset of patients with prolonged survival who had previously gone undetected when using traditional evaluations of treatment responsiveness.

## Discussion

MR images are central in the management of brain tumors, and they contain a wealth of information beyond what is perceptible by the human eye.^24^ High-dimensional imaging data can be leveraged to derive signatures that have quantifiable associations to biological features or clinical endpoints.^25–29^ This is particularly valuable in the context of glioblastoma, where intratumoral heterogeneity and ongoing genetic evolution hinder our ability to assess a tumor’s biological state after the initial surgical diagnosis. Here we report the results of our sex-specific radiomics model applied to an early phase 1 dendritic cell vaccine clinical trial. Most patients had an increase in systemic CD8^+^ T-cells over the course of treatment as determined by peripheral blood samples^23^, yet the median survival time was on par with the disease’s natural history.^1,2^ Spatiotemporal radiomics patterning identified a subset of these patients, those with a regional decrease of predicted T-cells in the T1Gd abnormal region with a simultaneous increase in the T2/FLAIR region, who seemed to be vaccine-responsive and had a median survival of 27 months. Our results illustrate several important points about the changing landscape of clinical trial assessment and radiomics best practices.

This study is the first, to our knowledge, to use image-localized biopsies in the voxelwise prediction of GBM microenvironmental biology from imaging. This approach addresses the challenge of GBM’s profound inter- and intratumoral heterogeneity. A biopsy from the edge of a GBM may have a completely different genotype, imaging phenotype, and therapeutic sensitivity compared to the tumor’s core.^30^ Without knowing the exact location from which the sample was harvested, biological and radiographic data can not be aligned in any meaningful way. Our image-localized samples allowed us to capture a wide spectrum of genotypes and imaging phenotypes within and between patients, thereby improving the specificity of our bioimaging relationships as well as the generalizability of our models.

Rather than combine all samples to build a single model that predicts T-cell abundance from imaging features, we trained separate male- and female-specific models. There are innumerable sex differences in cancer biology, and analyzing male and female data separately can reveal signals that are lost when analyzing all patients together.^31^ This oversight could hinder our ability to understand the biology of disease in its totality. In support of a sex-specific approach, our models outperformed the sex-agnostic model on unseen test data. Additionally, the most important imaging features to each model were derived from different sequences (T2 in males and T1Gd in females). It is particularly interesting that the most important feature in the sex-combined model was T2-related, as our data has a slight overrepresentation of male samples. These findings further support the idea that male and female samples should be analyzed separately so as not to overlook biologically relevant sex differences, and that sex-specific approaches should be considered by all who are building image-based models.

A spatiotemporal analysis of T-cell predictions in the dendritic cell vaccine clinical trial revealed a subset of patients with prolonged survival who had not previously been identified. Specifically, those whose predicted T-cells decreased within the T1Gd region while simultaneously increasing in the T2/FLAIR region had prolonged survival, even when adjusting for patient demographics and tumor genetics. Perhaps the most striking detail of this result is the timeframe over which the images were acquired. “Pre-treatment” imaging was taken immediately before the first vaccine dose, and “post-treatment” imaging was taken two months thereafter, while all patients were still alive. This spatial radiomics pattern was an early, measurable biomarker of treatment responsiveness that could theoretically prospectively aid physicians and patients in treatment decision-making. Further, these results challenge traditional methods of clinical trial assessment and suggest that evaluations of median survival across a diverse cohort may inadvertently overlook subpopulations who are treatment responsive.

The relationship between prolonged survival and divergent T-cell dynamics within T1Gd and T2/FLAIR ROIs could be explained by the underlying biology that gives rise to hyperintensity in each MRI sequence. T1Gd hyperintensity represents contrast extravasation into the brain parenchyma, which is suggestive of BBB disruption. The intact BBB selectively allows activated, but not naïve, T-cells to enter the brain.^32^ Therefore, accumulation of T-cells in the T1Gd ROI can occur secondary to BBB disruption and may not directly correlate with T-cell recruitment or activation. This has been shown in the context of focused ultrasound, a method by which the BBB is disrupted for enhanced regional delivery of drugs and cytokines. Following focused ultrasound, Chen et al.^33^ observed an increase in tumor-infiltrating lymphocytes. On the other hand, T2 hyperintensity represents increased water content and edema, which is often secondary to inflammation. Therefore, an increase in T-cells within this region is more likely to represent an active immune response. Taken together, T-cells decreasing within the T1Gd ROI and simultaneously increasing within the T2/FLAIR ROI could represent the activation and outward mobilization of immune effectors from a previously inactive contrast-enhancing niche that accumulated simply due to BBB disruption.

The strengths of both the clinical trial as well as our image-localized biopsy collection are the prospective collection of data using standardized protocols. Still, we acknowledge that there are limitations in the interpretation of our results. Intraoperative surgical navigation has a minimal but non-zero margin of error. This means that a tumor sample may have been harvested from a slightly different location than the coordinate which is recorded and used for downstream analyses. To address this, imaging features were calculated from a 5mm spherical region centered at the biopsy coordinate. Additionally, MR images were manually segmented and processed by a team of trained individuals. While this was performed under the close supervision of an experienced neuroradiologist, the manual nature of this pipeline introduces the potential for interpatient variation. Lastly, the use of CIBERSORT for the enumeration of T-cells has been robustly benchmarked and validated.^21,34^ Still, RNA sequencing deconvolution is an indirect estimation of immune cell abundance, and future studies could employ direct measures (e.g. immunohistochemistry, flow cytometry, single cell RNA sequencing) for the training of image-based models.

In conclusion, image-localized biopsies allow for the training of machine learning models that predict biological information from MRI. This has immense value in the context of tumors like glioblastoma, where there is spatial and temporal heterogeneity that cannot be assessed regularly or completely with tissue sampling. The utility of this method is highlighted in the context of the dendritic cell vaccine clinical trial, where patients on average did not have a survival benefit from the experimental therapy. Leveraging sex-specific T-cell radiomics models applied across multiple time points, we found a spatial radiomics pattern with a relationship to prolonged survival. In a field where hundreds of experimental therapies have been deemed “failures”, radiomics may be the key to improving our assessment of clinical trials and moving closer towards a reality of individualized medicine.

## Data Availability

All data produced in the present study are available upon reasonable request to the authors and/or are provided in supplementary Tables attached within the manuscript.

## Acknowledgements

The authors gratefully acknowledge the funding that made this research possible from the NIH (KRS: U54CA143970, U54CA193489, R01CA16437, KRS, NLT, LSH: U01CA220378; KRS, LSH, PC: U01CA250481), the NSF (KRS: 1903135), the Zicarelli Foundation (KRS), the James S. McDonnell Foundation (KRS, PC), the Ben and Catherine Ivy Foundation (KRS) and Mayo Clinic. We are also grateful to all of those who have contributed to elements of this work, particularly current and past members of the image analysis teams and the glioma biopsy protocol teams, including: Barrett Anderies, Jessica Bauer, Spencer Bayless, Hend Bcharach, Regina Becker, Sameer Channer, Brenden Doyle, Lysette Elsner, Lily Esaleh, Ashlyn Gonzales, Crystal Harris, Morgan Hatlestead, Ryan Hess, Sandra Johnston, Yvette Lassiter-Morris, Julia Lorence, Ashley Napier, Ashley Nespodzany, Lisa Paulson, Cassandra Rickertsen, Sejal Shanbhag, Sarah Van Dijk, and Scott Whitmire.

