## Supplemental Table 5 for "Image-based models of T-cell distribution identify a clinically meaningful response to a dendritic cell vaccine in patients with glioblastoma"

| **Criteria** | |  |
| --- | --- | --- |
| 1 | Image protocol quality - well-documented image protocols (for example, contrast, slice thickness, energy, etc.) and/or usage of public image protocols allow reproducibility/replicability | 1 |
| 2 | Multiple segmentations - possible actions are: segmentation by different physicians/algorithms/software, perturbing segmentations by (random) noise, segmentation at different breathing cycles. Analyse feature robustness to segmentation variabilities | 1 |
| 3 | Phantom study on all scanners - detect inter-scanner differences and vendor-dependent features. Analyse feature robustness to these sources of variability | 0 |
| 4 | Imaging at multiple time points - collect images of individuals at additional time points. Analyse feature robustness to temporal variabilities (for example, organ movement, organ expansion/shrinkage) | 1 |
| 5 | Feature reduction or adjustment for multiple testing - decreases the risk of overfitting. Overfitting is inevitable if the number of features exceeds the number of samples. Consider feature robustness when selecting features | 3 |
| 6 | Multivariable analysis with non radiomics features (for example, EGFR mutation) - is expected to provide a more holistic model. Permits correlating/inferencing between radiomics and non radiomics features | 1 |
| 7 | Detect and discuss biological correlates - demonstration of phenotypic differences (possibly associated with underlying gene–protein expression patterns) deepens understanding of radiomics and biology | 1 |
| 8 | Cut-off analyses - determine risk groups by either the median, a previously published cut-off or report a continuous risk variable. Reduces the risk of reporting overly optimistic results | 1 |
| 9 | Discrimination statistics - report discrimination statistics (for example, C-statistic, ROC curve, AUC) and their statistical significance (for example, p-values, confidence intervals). One can also apply resampling method (for example, bootstrapping, cross-validation) | 0 |
| 10 | Calibration statistics - report calibration statistics (for example, Calibration-in-the-large/slope, calibration plots) and their statistical significance (for example, *P*-values, confidence intervals). One can also apply resampling method (for example, bootstrapping, cross-validation) | 0 |
| 11 | Prospective study registered in a trial database - provides the highest level of evidence supporting the clinical validity and usefulness of the radiomics biomarker | 7 |
| 12 | Validation - the validation is performed without retraining and without adaptation of the cut-off value, provides crucial information with regard to credible clinical performance | 2 |
| 13 | Comparison to 'gold standard' - assess the extent to which the model agrees with/is superior to the current 'gold standard' method (for example, TNM-staging for survival prediction). This comparison shows the added value of radiomics | 2 |
| 14 | Potential clinical utility - report on the current and potential application of the model in a clinical setting (for example, decision curve analysis). | 2 |
| 15 | Cost-effectiveness analysis - report on the cost-effectiveness of the clinical application (for example, QALYs generated) | 0 |
| 16 | Open science and data - make code and data publicly available. Open science facilitates knowledge transfer and reproducibility of the study | 0 |
|  | Total: 22 |  |
