## Supplemental Table 1 for "Image-based models of T-cell distribution identify a clinically meaningful response to a dendritic cell vaccine in patients with glioblastoma"

| BiopsyPatientID | Status | Sex | Age Range | Number of Samples | Contrast Enhancing |
|---|---|---|---|---|---|
| 1 | Recurrent | F | 20-24 | 3 | 0 |
| 2 | Recurrent | F | 50-54 | 2 | 2 |
| 3 | Primary | M | 65-69 | 1 | 0 |
| 4 | Primary | F | 55-59 | 9 | 7 |
| 5 | Primary | M | 70-74 | 7 | 2 |
| 6 | Recurrent | F | 25-29 | 4 | 0 |
| 7 | Recurrent | F | 40-44 | 4 | 0 |
| 8 | Recurrent | M | 60-64 | 2 | 0 |
| 9 | Recurrent | F | 60-64 | 5 | 4 |
| 10 | Primary | M | 70-74 | 3 | 2 |
| 11 | Primary | M | 30-34 | 3 | 1 |
| 12 | Primary | M | 65-70 | 4 | 0 |
| 13 | Primary | F | 55-59 | 2 | 0 |
| 14 | Primary | M | 45-49 | 6 | 2 |
| 15 | Primary | F | 50-54 | 2 | 2 |
| 16 | Primary | M | 65-69 | 3 | 2 |
| 17 | Recurrent | M | 25-29 | 3 | 0 |
| 18 | Recurrent | M | 20-24 | 3 | 0 |
| 19 | Primary | F | 25-29 | 1 | 1 |
| 20 | Primary | M | 70-74 | 7 | 4 |
| 21 | Primary | M | 60-64 | 4 | 2 |
| 22 | Primary | M | 55-59 | 3 | 2 |
| 23 | Recurrent | F | 70-74 | 2 | 0 |
| 24 | Primary | F | 45-49 | 7 | 2 |
| 25 | Primary | M | 70-74 | 2 | 2 |
| 26 | Recurrent | M | 55-59 | 3 | 0 |
| 27 | Primary | M | 65-69 | 3 | 0 |
| 28 | Primary | M | 70-74 | 4 | 2 |
| 29 | Recurrent | M | 40-44 | 3 | 2 |
| 30 | Recurrent | F | 60-64 | 6 | 5 |
| 31 | Primary | F | 55-59 | 7 | 2 |
| 32 | Primary | F | 25-29 | 6 | 1 |
| 33 | Primary | F | 45-49 | 2 | 0 |
| 34 | Primary | M | 55-59 | 2 | 0 |
| 35 | Primary | F | 70-74 | 1 | 0 |
| 36 | Recurrent | M | 30-34 | 1 | 1 |
| 37 | Primary | M | 65-69 | 4 | 0 |
| 38 | Primary | M | 55-59 | 2 | 2 |
| 39 | Primary | M | 60-64 | 1 | 0 |
| 40 | Primary | M | 20-24 | 2 | 1 |
| 41 | Primary | F | 25-29 | 3 | 2 |
| 42 | Primary | M | 40-44 | 2 | 2 |

| | | | | | |
|---|---|---|---|---|---|
| 43 | Primary | M | 75-79 | 3 | 0 |

| Non-Enhancing | Necrosis |
|---|---|
| 2 | 1 |
| 0 | 0 |
| 1 | 0 |
| 2 | 0 |
| 5 | 0 |
| 4 | 0 |
| 3 | 1 |
| 1 | 1 |
| 1 | 0 |
| 1 | 0 |
| 2 | 0 |
| 4 | 0 |
| 2 | 0 |
| 4 | 0 |
| 0 | 0 |
| 1 | 0 |
| 1 | 2 |
| 3 | 0 |
| 0 | 0 |
| 3 | 0 |
| 2 | 0 |
| 1 | 0 |
| 2 | 0 |
| 4 | 1 |
| 0 | 0 |
| 3 | 0 |
| 3 | 0 |
| 1 | 1 |
| 0 | 1 |
| 1 | 0 |
| 5 | 0 |
| 5 | 0 |
| 2 | 0 |
| 1 | 1 |
| 0 | 1 |
| 0 | 0 |
| 4 | 0 |
| 0 | 0 |
| 0 | 1 |
| 1 | 0 |
| 1 | 0 |
| 0 | 0 |

1 2

| BiopsyPatientID | SampleID | Enhancing |
|---|---|---|
| 1 | 1 | 2 |
| 1 | 2 | 1 |
| 1 | 3 | 1 |
| 2 | 1 | 0 |
| 2 | 2 | 0 |
| 3 | 1 | 1 |
| 4 | 1 | 0 |
| 4 | 2 | 0 |
| 4 | 3 | 0 |
| 4 | 4 | 0 |
| 4 | 5 | 0 |
| 4 | 6 | 0 |
| 4 | 7 | 0 |
| 4 | 8 | 1 |
| 4 | 9 | 1 |
| 5 | 1 | 0 |
| 5 | 2 | 1 |
| 5 | 3 | 0 |
| 5 | 4 | 1 |
| 5 | 5 | 1 |
| 5 | 6 | 1 |
| 5 | 7 | 1 |
| 6 | 1 | 1 |
| 6 | 2 | 1 |
| 6 | 3 | 1 |
| 6 | 4 | 1 |
| 7 | 1 | 2 |
| 7 | 2 | 1 |
| 7 | 3 | 1 |
| 7 | 4 | 1 |
| 8 | 1 | 1 |
| 8 | 2 | 2 |
| 9 | 1 | 1 |
| 9 | 2 | 0 |
| 9 | 3 | 0 |
| 9 | 4 | 0 |
| 9 | 5 | 0 |
| 10 | 1 | 0 |
| 10 | 2 | 0 |
| 10 | 3 | 1 |
| 11 | 1 | 0 |
| 11 | 2 | 1 |

| | | |
|---|---|---|
| 11 | 3 | 1 |
| 12 | 1 | 1 |
| 12 | 2 | 1 |
| 12 | 3 | 1 |
| 12 | 4 | 1 |
| 13 | 1 | 1 |
| 13 | 2 | 1 |
| 14 | 1 | 1 |
| 14 | 2 | 1 |
| 14 | 3 | 0 |
| 14 | 4 | 1 |
| 14 | 5 | 0 |
| 14 | 6 | 1 |
| 15 | 1 | 0 |
| 15 | 2 | 0 |
| 16 | 1 | 0 |
| 16 | 2 | 1 |
| 16 | 3 | 0 |
| 17 | 1 | 2 |
| 17 | 2 | 2 |
| 17 | 3 | 1 |
| 18 | 1 | 1 |
| 18 | 2 | 1 |
| 18 | 3 | 1 |
| 19 | 1 | 0 |
| 20 | 1 | 1 |
| 20 | 2 | 1 |
| 20 | 3 | 0 |
| 20 | 1 | 0 |
| 20 | 2 | 1 |
| 20 | 3 | 0 |
| 20 | 4 | 0 |
| 21 | 1 | 1 |
| 21 | 2 | 1 |
| 21 | 3 | 0 |
| 21 | 4 | 0 |
| 22 | 1 | 1 |
| 22 | 2 | 0 |
| 22 | 3 | 0 |
| 23 | 1 | 1 |
| 23 | 2 | 1 |
| 24 | 1 | 1 |
| 24 | 2 | 1 |

| | | |
|---|---|---|
| 24 | 3 | 1 |
| 24 | 4 | 1 |
| 24 | 5 | 2 |
| 24 | 6 | 0 |
| 24 | 7 | 0 |
| 25 | 1 | 0 |
| 25 | 2 | 0 |
| 26 | 1 | 1 |
| 26 | 2 | 1 |
| 26 | 3 | 1 |
| 27 | 1 | 1 |
| 27 | 2 | 1 |
| 27 | 3 | 1 |
| 28 | 1 | 0 |
| 28 | 2 | 0 |
| 28 | 3 | 2 |
| 28 | 4 | 1 |
| 29 | 1 | 0 |
| 29 | 2 | 0 |
| 29 | 3 | 2x |
| 30 | 1 | 1 |
| 30 | 2 | 0 |
| 30 | 3 | 0 |
| 30 | 4 | 0 |
| 30 | 5 | 0 |
| 30 | 6 | 0 |
| 31 | 1 | 1 |
| 31 | 2 | 1 |
| 31 | 3 | 1 |
| 31 | 4 | 1 |
| 31 | 5 | 1 |
| 31 | 6 | 0 |
| 31 | 7 | 0 |
| 32 | 1 | 0 |
| 32 | 2 | 1 |
| 32 | 3 | 1 |
| 32 | 4 | 1 |
| 32 | 5 | 1 |
| 32 | 6 | 1 |
| 33 | 1 | 1 |
| 33 | 2 | 1 |
| 34 | 1 | 1 |
| 34 | 2 | 2 |

| | | |
|---|---|---|
| 35 | 1 | 2 |
| 36 | 1 | 0 |
| 37 | 1 | 1 |
| 37 | 2 | 1 |
| 37 | 3 | 1 |
| 37 | 4 | 1 |
| 38 | 1 | 0 |
| 38 | 2 | 0 |
| 39 | 1 | 2 |
| 40 | 1 | 0 |
| 40 | 2 | 1 |
| 41 | 1 | 0 |
| 41 | 2 | 0 |
| 41 | 3 | 1 |
| 42 | 1 | 0 |
| 42 | 2 | 0 |
| 43 | 1 | 2 |
| 43 | 2 | 2 |
| 43 | 3 | 1 |
