## Supplemental Table 2 for "Image-based models of T-cell distribution identify a clinically meaningful response to a dendritic cell vaccine in patients with glioblastoma"

| PatientID | SampleID | B cells naive | B cells mem | Plasma cells | T cells CD8 | T cells CD4 n |
|---|---|---|---|---|---|---|
| 1 | 1 | 0.00598982 | 0.01262255 | 0.0051959 | 0.08420722 | 0 |
| 1 | 2 | 0.02984128 | 0.00498057 | 0 | 0.0394492 | 0 |
| 1 | 3 | 0.01803806 | 0.01574034 | 0 | 0.05301682 | 0.03420226 |
| 6 | 1 | 0 | 0.09313093 | 0.01369827 | 0.10621282 | 0 |
| 6 | 2 | 0.02911475 | 0.00377315 | 0 | 0.10509365 | 0 |
| 6 | 3 | 0 | 0.07485492 | 0.0066624 | 0.08260227 | 0 |
| 6 | 4 | 0.03940507 | 0.00885998 | 0.00015042 | 0.08123485 | 0 |
| 7 | 1 | 0.0642469 | 0 | 0.00224362 | 0.03197727 | 0 |
| 7 | 2 | 0.03138916 | 0 | 0 | 0.60832212 | 0 |
| 7 | 3 | 0 | 0.08963757 | 0.0153238 | 0.08710857 | 0 |
| 7 | 4 | 0.0536154 | 0 | 0.0016685 | 0.07106514 | 0 |
| 8 | 1 | 0.03444752 | 0 | 0 | 0 | 0 |
| 8 | 2 | 0.03296278 | 0 | 0 | 0.05540629 | 0 |
| 9 | 1 | 0.04958525 | 0.02647949 | 0.02093393 | 0.16711044 | 0 |
| 9 | 2 | 0.01287622 | 0.02276703 | 0 | 0.13282642 | 0 |
| 9 | 3 | 0.00874198 | 0.05909947 | 0 | 0.15357607 | 0 |
| 9 | 4 | 0 | 0.06248179 | 0 | 0.13326406 | 0 |
| 9 | 5 | 0.02188332 | 0.03619812 | 0 | 0.07881265 | 0 |
| 2 | 1 | 0.06693688 | 0.04820609 | 0.12745952 | 0.21855948 | 0 |
| 2 | 2 | 0.15146035 | 0.02882192 | 0.08920116 | 0.2455968 | 0 |
| 4 | 1 | 0.03555916 | 0 | 0 | 0.0426012 | 0 |
| 4 | 2 | 0.01047943 | 0.01438218 | 0 | 0.06298313 | 0 |
| 4 | 3 | 0.07031703 | 0 | 0 | 0.07205641 | 0.03631455 |
| 4 | 4 | 0.03544193 | 0.00811438 | 0 | 0.06406698 | 0 |
| 4 | 5 | 0.03610727 | 0.00633357 | 0 | 0.0522223 | 0.00796603 |
| 4 | 6 | 0.02974733 | 0.02405064 | 0 | 0.06536318 | 0 |
| 4 | 7 | 0.02722737 | 0 | 0 | 0.03003962 | 0.00287164 |
| 4 | 8 | 0.02582621 | 0.01521293 | 0 | 0.08179037 | 0 |
| 4 | 9 | 0.04730382 | 0 | 0 | 0.05309488 | 0 |
| 5 | 1 | 0.0334791 | 0 | 0.04571365 | 0.21593659 | 0 |
| 5 | 2 | 0.00489849 | 0.01515155 | 0.0112282 | 0.37013488 | 0 |
| 5 | 3 | 0.01639095 | 0.00918549 | 0.00922947 | 0.16873847 | 0 |
| 5 | 4 | 0.02991327 | 0.01782051 | 0 | 0.18624899 | 0 |
| 5 | 5 | 0.19081316 | 0 | 0.07299796 | 0.31133107 | 0 |
| 5 | 6 | 0.05040295 | 0.01019419 | 0.03874937 | 0.09731327 | 0 |
| 5 | 7 | 0.00278297 | 0.02614738 | 0.02550253 | 0.3575199 | 0 |
| 10 | 1 | 0.01397315 | 0.02314109 | 0.00491278 | 0.05580585 | 0 |
| 10 | 2 | 0 | 0.13831637 | 0.083939 | 0.19116589 | 0 |
| 10 | 3 | 0.05963724 | 0 | 0.05905052 | 0.31685445 | 0 |
| 11 | 1 | 0.03052835 | 0.02096379 | 0.01440774 | 0.04251249 | 0 |
| 11 | 2 | 0.05194042 | 0 | 0.00662427 | 0.05796517 | 0 |
| 11 | 3 | 0.01038001 | 0.06972856 | 0 | 0.01799265 | 0 |

| | | | | | | |
|---|---|---|---|---|---|---|
| 12 | 1 | 0.01182675 | 0.02072644 | 0 | 0.10552158 | 0 |
| 12 | 2 | 0.00278466 | 0.01683826 | 0 | 0.29604166 | 0 |
| 12 | 3 | 0.0373671 | 0.00441186 | 0 | 0.28467722 | 0 |
| 12 | 4 | 0.02956175 | 0.0009209 | 0 | 0.15822377 | 0 |
| 13 | 1 | 0 | 0.05359195 | 0.00050818 | 0.10846108 | 0 |
| 13 | 2 | 0 | 0.07589255 | 0.01726855 | 0.15833947 | 0 |
| 17 | 1 | 0 | 0.11485003 | 0.02536787 | 0.1499063 | 0 |
| 19 | 1 | 0.0063652 | 0.03640454 | 0.01165744 | 0.07757031 | 0 |
| 24 | 3 | 0 | 0.00841505 | 0 | 0 | 0 |
| 28 | 1 | 0.043905 | 0 | 0.04581195 | 0.20172654 | 0 |
| 31 | 4 | 0 | 0.03590333 | 0.02904404 | 0.07399549 | 0 |
| 14 | 5 | 0 | 0.05095754 | 0.01479061 | 0.08121324 | 0 |
| 20 | 2 | 0 | 0.0795779 | 0.00687522 | 0.04191504 | 0 |
| 17 | 2 | 0 | 0.08730662 | 0.031187 | 0.07508459 | 0 |
| 22 | 1 | 0.05593644 | 0.04410074 | 0.0457929 | 0.52026885 | 0 |
| 24 | 4 | 0 | 0.05883282 | 0.01829659 | 0.03443358 | 0 |
| 28 | 2 | 0.02501534 | 0.02405993 | 0.03519273 | 0.44606516 | 0 |
| 31 | 5 | 0 | 0.05930606 | 0.00961266 | 0.07711337 | 0 |
| 14 | 6 | 0.01143264 | 0.05223439 | 0.00487959 | 0.23473966 | 0 |
| 20 | 3 | 0.08660835 | 0 | 0 | 0.02726288 | 0.02529288 |
| 17 | 3 | 0.00589394 | 0 | 0 | 0.22480469 | 0 |
| 22 | 2 | 0.08834308 | 0.02927906 | 0.06159612 | 0.79736945 | 0 |
| 24 | 5 | 0 | 0.0933229 | 0.00674467 | 0.07473001 | 0 |
| 28 | 3 | 0.0317948 | 0.00186902 | 0.1060461 | 0.22005514 | 0 |
| 31 | 6 | 0 | 0.1394534 | 0.01636624 | 0 | 0 |
| 15 | 1 | 0.13721101 | 0 | 0.0769363 | 0.12066174 | 0 |
| 20 | 1 | 0.01304146 | 0.00964483 | 0 | 0.21049929 | 0 |
| 22 | 3 | 0.07790967 | 0.13746425 | 0.03467957 | 0.41400062 | 0 |
| 24 | 6 | 0 | 0.08728486 | 0.00767308 | 0.08685758 | 0 |
| 28 | 4 | 0.05140777 | 0 | 0.04165242 | 0.38298544 | 0 |
| 31 | 7 | 0.0607566 | 0.04112952 | 0 | 0.02093895 | 0 |
| 15 | 2 | 0.10911977 | 0.0352329 | 0.1069346 | 0.16123562 | 0 |
| 21 | 1 | 0.05134846 | 0 | 0 | 0 | 0 |
| 25 | 1 | 0.06033377 | 0 | 0 | 0 | 0 |
| 24 | 7 | 0.02086934 | 0 | 0 | 0.08625717 | 0 |
| 14 | 1 | 0.00766211 | 0.01901665 | 0.02423641 | 0.05183085 | 0 |
| 16 | 1 | 0.00518117 | 0.00372331 | 0.00665114 | 0.10131971 | 0 |
| 20 | 2 | 0.05444821 | 0 | 0 | 0.0594002 | 0 |
| 21 | 2 | 0.03682706 | 0.01361374 | 0 | 0.06951671 | 0 |
| 25 | 2 | 0.02160542 | 0 | 0 | 0 | 0 |
| 27 | 1 | 0.03138677 | 0.00566843 | 0.05316636 | 0 | 0 |
| 31 | 1 | 0.01705907 | 0.04050347 | 0.00333385 | 0.02829925 | 0 |
| 14 | 2 | 0 | 0.15606821 | 0.03282 | 0.37373018 | 0 |

| | | | | | | |
|---|---|---|---|---|---|---|
| 16 | 2 | 0 | 0.03944267 | 0.02652586 | 0.1993671 | 0 |
| 20 | 3 | 0.00906358 | 0.03806973 | 0.00242945 | 0.16331987 | 0 |
| 21 | 3 | 0.03408927 | 0.01940398 | 0 | 0 | 0.0125292 |
| 24 | 1 | 0.02561919 | 0.00603252 | 0 | 0.0510246 | 0 |
| 27 | 2 | 0.07717656 | 0 | 0.00166665 | 0.05598946 | 0 |
| 31 | 2 | 0.01917336 | 0.00463584 | 0.02827813 | 0.15278089 | 0 |
| 14 | 3 | 0 | 0.01146865 | 0.03161599 | 0.27616644 | 0 |
| 16 | 3 | 0.0298151 | 0.00186389 | 0 | 0.11579486 | 0 |
| 20 | 4 | 0.04446192 | 0.00304398 | 0 | 0.18347125 | 0 |
| 21 | 4 | 0.0426006 | 0 | 0 | 0 | 0.00306378 |
| 24 | 2 | 0.03875689 | 0.01216395 | 0 | 0.07676154 | 0 |
| 27 | 3 | 0.02480928 | 0.04084358 | 0 | 0.0725178 | 0 |
| 31 | 3 | 0.02288622 | 0.08538803 | 0.02030102 | 0.11439899 | 0 |
| 14 | 4 | 0.03191147 | 0 | 0.01020172 | 0.12521081 | 0 |
| 20 | 1 | 0.07535458 | 0 | 0.00108385 | 0.10659124 | 0 |
| 18 | 1 | 0.03768919 | 0.0081086 | 0 | 0.07617899 | 0 |
| 18 | 2 | 0.01476075 | 0.00643773 | 0 | 0.12463467 | 0 |
| 18 | 3 | 0.03358432 | 0 | 0 | 0.04075283 | 0 |
| 23 | 1 | 0.06160439 | 0 | 0 | 0.11861035 | 0 |
| 23 | 2 | 0.09738532 | 0 | 0 | 0.12337003 | 0.0478303 |
| 29 | 1 | 0.1096615 | 0 | 0.06279284 | 0.51832396 | 0 |
| 29 | 2 | 0.08358646 | 0 | 0.03898111 | 0.69559408 | 0 |
| 29 | 3 | 0.04358499 | 0 | 0.03153109 | 0.27250284 | 0 |
| 30 | 1 | 0 | 0.13764635 | 0 | 0.07460066 | 0 |
| 30 | 2 | 0.04328486 | 0 | 0 | 0.02734924 | 0 |
| 30 | 3 | 0.01535104 | 0.01489769 | 0.0014873 | 0.1130412 | 0 |
| 30 | 4 | 0.03386468 | 0.06157222 | 0.03880859 | 0.07428523 | 0 |
| 30 | 5 | 0 | 0.11156132 | 0 | 0.15862112 | 0 |
| 30 | 6 | 0.06875574 | 0.07586753 | 0.0194337 | 0.03223863 | 0 |
| 32 | 1 | 0.05247529 | 0 | 0.00164769 | 0.05511914 | 0 |
| 32 | 2 | 0.03338136 | 0.01480165 | 0.0023898 | 0.15410658 | 0 |
| 32 | 3 | 0.04125927 | 0 | 0.0191921 | 0.06984998 | 0 |
| 32 | 4 | 0.03571994 | 0.0057677 | 0 | 0.12080114 | 0 |
| 32 | 5 | 0.03953165 | 0.00425115 | 0.00652587 | 0.09803533 | 0 |
| 32 | 6 | 0.06362201 | 0 | 0.00386161 | 0.1804221 | 0 |
| 33 | 1 | 0.25669244 | 0 | 0.08175174 | 0.60291336 | 0 |
| 33 | 2 | 0.02658436 | 0 | 0.02021615 | 0.04118159 | 0 |
| 34 | 1 | 0.09157136 | 0 | 0.00260155 | 0.18148846 | 0 |
| 34 | 2 | 0.02803104 | 0.0592542 | 0 | 0.15902125 | 0 |
| 35 | 1 | 0.00111847 | 0.01624074 | 0.00365412 | 0.20862612 | 0 |
| 36 | 1 | 0.01653962 | 0.07115528 | 0.03427177 | 0.14238134 | 0 |
| 37 | 1 | 0.02971902 | 0 | 0 | 0.03537915 | 0.0144595 |
| 37 | 2 | 0 | 0.05014085 | 0.00104838 | 0.03026178 | 0.04324519 |

| | | | | | | |
|---|---|---|---|---|---|---|
| 37 | 3 | 0.02311681 | 0.00072455 | 0 | 0.05939647 | 0.01336638 |
| 37 | 4 | 0 | 0.06972402 | 0 | 0.07824058 | 0 |
| 38 | 1 | 0.02686812 | 0.02530681 | 0 | 0.04319089 | 0.02837598 |
| 38 | 2 | 0.02228933 | 0.0386598 | 0 | 0.08197437 | 0 |
| 39 | 1 | 0.01713138 | 0.01021676 | 0 | 0.07024341 | 0 |
| 40 | 1 | 0.00416649 | 0.05205199 | 0 | 0.02102613 | 0.01511571 |
| 40 | 2 | 0 | 0.02977106 | 0.01108345 | 0.01794977 | 0 |
| 41 | 1 | 0.1247653 | 0 | 0.21137195 | 0.15703698 | 0 |
| 41 | 2 | 0.04298144 | 0.03808717 | 0.04564225 | 0.10398407 | 0 |
| 41 | 3 | 0.01431643 | 0.08749005 | 0 | 0.04364234 | 0 |
| 42 | 1 | 0.12794139 | 0 | 0 | 0.11896389 | 0 |
| 42 | 2 | 0.0566986 | 0.01440451 | 0 | 0.09714278 | 0 |
| 43 | 1 | 0.05052021 | 0 | 0.04338775 | 0.04551798 | 0 |
| 43 | 2 | 0.00311587 | 0.02791022 | 0.00509225 | 0.37192092 | 0 |
| 43 | 3 | 0.00754492 | 0.02896655 | 0.00590056 | 0.04511615 | 0 |
| 26 | 1 | 0 | 0.05283948 | 0.01545494 | 0.19305394 | 0 |
| 26 | 2 | 0.01633066 | 0 | 0.01429098 | 0.53291211 | 0 |
| 26 | 3 | 0 | 0.09819004 | 0.00386127 | 0.13848465 | 0 |

| T cells CD4 n | T cells CD4 n | T cells follicu | T cells regula | T cells gamm | NK cells resti | NK cells activ |
|---|---|---|---|---|---|---|
| 0.15456774 | 0 | 0.14628516 | 0.00251441 | 0 | 0.01792994 | 0.12688174 |
| 0.01868145 | 0 | 0.07617277 | 0.05906373 | 0 | 0.09415208 | 0 |
| 0.05074779 | 0 | 0.02111511 | 0.03086017 | 0 | 0.05821667 | 0 |
| 0.10081106 | 0.01863744 | 0.02111435 | 0.00418455 | 0 | 0.03877205 | 0.01887548 |
| 0.05350956 | 0 | 0.05924141 | 0.02554562 | 0.0103975 | 0.02646059 | 0.01714182 |
| 0.15006116 | 0.04999165 | 0.02482658 | 0 | 0 | 0.00270575 | 0.04580799 |
| 0.22113155 | 0 | 0 | 0.03232965 | 0 | 0.07082238 | 0 |
| 0.12996084 | 0 | 0.06684431 | 0.01325802 | 0 | 0.0628084 | 0.03314621 |
| 0.07242702 | 0.04205488 | 0.15218512 | 0.1293594 | 0.04919901 | 0.0371139 | 0 |
| 0 | 0 | 0.0928996 | 0.07151387 | 0.03098632 | 0.08478046 | 0.04950622 |
| 0.1934491 | 0.03069286 | 0.03470965 | 0.05265678 | 0 | 0.01162695 | 0.04220487 |
| 0.21283754 | 0.00162271 | 0.01776861 | 0.00541 | 0 | 0.07201327 | 0 |
| 0.11760714 | 0 | 0.03084892 | 0.00317321 | 0 | 0 | 0.11419338 |
| 1.09251613 | 0 | 0 | 0.08752466 | 0 | 0 | 0.17823749 |
| 0.0746964 | 0.04489748 | 0.01922901 | 0 | 0 | 0 | 0.08558366 |
| 0.40244686 | 0.13996088 | 0.01107817 | 0.04950411 | 0 | 0.01004288 | 0.08761094 |
| 0.32280179 | 0.0092765 | 0.0193454 | 0.06917979 | 0.01371391 | 0 | 0.1773414 |
| 0.35453852 | 0 | 0.02443223 | 0.06440069 | 0 | 0 | 0.10486733 |
| 0.06168055 | 0 | 0.08017418 | 0.0563534 | 0 | 0 | 0.14028606 |
| 0.06371617 | 0.01876336 | 0.09143795 | 0.13054419 | 0 | 0 | 0.15917641 |
| 0.26596638 | 0 | 0.05230372 | 0 | 0 | 0.01620079 | 0.0936823 |
| 0.18103461 | 0 | 0.06430471 | 0.02117511 | 0 | 0.00993409 | 0.06912913 |
| 0.11535899 | 0.02529466 | 0.01958164 | 0 | 0 | 0.04785201 | 0.05469582 |
| 0.34312329 | 0 | 0.07779718 | 0.02057622 | 0 | 0.01342135 | 0.16910779 |
| 0.23800143 | 0.22278259 | 0.0195062 | 0 | 0 | 0.06785568 | 0.07545823 |
| 0.06857678 | 0.01247303 | 0.02792632 | 0.0825752 | 0 | 0.05752091 | 0.10224995 |
| 0.2306671 | 0.05385544 | 0.00707078 | 0 | 0 | 0.10847972 | 0.01110741 |
| 0.43946945 | 0 | 0.04839166 | 0.04474726 | 0 | 0.06663668 | 0.07970898 |
| 0.12002254 | 0 | 0.06809134 | 0.01330307 | 0 | 0.05012846 | 0.05126646 |
| 0.41614709 | 0 | 0.07188845 | 0.0340244 | 0 | 0 | 0.03951817 |
| 0.10409405 | 0 | 0.02903703 | 0.12308765 | 0 | 0 | 0.17470172 |
| 0.28189746 | 0 | 0.04697462 | 0.00499067 | 0 | 0.06930125 | 0.01285508 |
| 0.11228848 | 0.0233923 | 0.07279139 | 0 | 0 | 0.08641903 | 0.01342536 |
| 0.4760004 | 0 | 0.07727418 | 0 | 0.1554551 | 0 | 0.19851232 |
| 0.08989526 | 0 | 0.12095353 | 0.07230031 | 0.18463359 | 0 | 0.18153534 |
| 0.51031852 | 0 | 0.06823715 | 0.11470083 | 0 | 0.05692501 | 0.04133876 |
| 0.1497489 | 0 | 0.07769133 | 0 | 0.06846276 | 0 | 0.13910984 |
| 0.34529883 | 0 | 0.04710787 | 0.06876094 | 0 | 0 | 0.34732698 |
| 0.12234957 | 0.01572967 | 0.27902061 | 0.37774778 | 0.00600399 | 0 | 0.21266543 |
| 0.23444129 | 0 | 0.03046153 | 0.02669702 | 0 | 0 | 0.09727433 |
| 0.16663032 | 0 | 0.04392198 | 0.03322188 | 0 | 0 | 0.13517971 |
| 0.11694127 | 0 | 0 | 0.0024698 | 0 | 0.05538097 | 0.0172043 |

| | | | | | | |
|---|---|---|---|---|---|---|
| 0.07809092 | 0 | 0.02150863 | 0.00094627 | 0 | 0.03188405 | 0.12251322 |
| 0.04580399 | 0 | 0.02276699 | 0.06210468 | 0 | 0 | 0.07022345 |
| 0.1495771 | 0 | 0.06521283 | 0.13700666 | 0 | 0 | 0.1032868 |
| 0.01566625 | 0 | 0.01748267 | 0.03728115 | 0 | 0.03352359 | 0.06655802 |
| 0.44058754 | 0 | 0.09001794 | 0.04682793 | 0 | 0.1672988 | 0.17190554 |
| 0.37517871 | 0 | 0.04213729 | 0.05248573 | 0 | 0.12531323 | 0.0103229 |
| 0.22402278 | 0 | 0.06374239 | 0.06241817 | 0 | 0 | 0.30059366 |
| 0.10527312 | 0.03662636 | 0 | 0.02830595 | 0 | 0.03590572 | 0.00355336 |
| 0.20471155 | 0 | 0.04266628 | 0 | 0.01402042 | 0.02355268 | 0.07061694 |
| 0.31849231 | 0 | 0.01504401 | 0.07813457 | 0.00900315 | 0.03597331 | 0 |
| 0.13600983 | 0.02134277 | 0.05106504 | 0 | 0.01881773 | 0.04509946 | 0.0063791 |
| 0.15923295 | 0.05113298 | 0 | 0 | 0.03095645 | 0 | 0.09827655 |
| 0.39573323 | 0.08605739 | 0 | 0.04426716 | 0 | 0.15040478 | 0 |
| 0.02232952 | 0.00515671 | 0.04429739 | 0.06561596 | 0.01000802 | 0.06292914 | 0 |
| 0.65285569 | 0 | 0.07120498 | 0.15633221 | 0.01675971 | 0 | 0.13470628 |
| 0.15708336 | 0 | 0.0306463 | 0.01675218 | 0 | 0.17646574 | 0 |
| 0.19002401 | 0 | 0.10894468 | 0.13001299 | 0 | 0.07276872 | 0 |
| 0.10513911 | 0 | 0.06236244 | 0.01233202 | 0.10401515 | 0 | 0.07425426 |
| 0.20361844 | 0 | 0.02428531 | 0.10347666 | 0 | 0.01337955 | 0.21209208 |
| 0.13252192 | 0 | 0.01537252 | 0.05870816 | 0 | 0.02091526 | 0.14668698 |
| 0.12688252 | 0 | 0.03663058 | 0.06127762 | 0 | 0.00824472 | 0.09365314 |
| 0.7375137 | 0 | 0.10466966 | 0.39867839 | 0 | 0.02934859 | 0.38963412 |
| 0.10430701 | 0 | 0.09144885 | 0.05426596 | 0 | 0.04168944 | 0.04721624 |
| 0.35613966 | 0.20228256 | 0.04625758 | 0 | 0 | 0.05887633 | 0.00070315 |
| 0.19092487 | 0.01148914 | 0.04693485 | 0 | 0.03001233 | 0 | 0.12580866 |
| 0.29531071 | 0 | 0.09158457 | 0.01368823 | 0 | 0 | 0.07809985 |
| 0.31108146 | 0 | 0.02836187 | 0.06534499 | 0 | 0.00708474 | 0.12093851 |
| 0.73106692 | 0 | 0.00631377 | 0.15360872 | 0.10161695 | 0 | 0.20358244 |
| 0.41392483 | 0 | 0.04137249 | 0.02828489 | 0 | 0.17405483 | 0.08301908 |
| 0 | 0.09277793 | 0.16239108 | 0.11801532 | 0.11131755 | 0.02562649 | 0 |
| 0.20056242 | 0.00553399 | 0 | 0.00371387 | 0 | 0.06676439 | 0 |
| 0.00883396 | 0 | 0.06666976 | 0.0877902 | 0 | 0 | 0.20784946 |
| 0.18990234 | 0 | 0.06437305 | 0.11326991 | 0 | 0.09279338 | 0.03066984 |
| 0.30085618 | 0 | 0.02991065 | 0 | 0 | 0 | 0.17425473 |
| 0.3476642 | 0 | 0.02851434 | 0.02014273 | 0 | 0.12125819 | 0.03427459 |
| 0.1590828 | 0 | 0.09778895 | 0.00761101 | 0.16072839 | 0 | 0.1940811 |
| 0.05790534 | 0 | 0.07588486 | 0.03331174 | 0 | 0 | 0.1070342 |
| 0.42064787 | 0.08872169 | 0.00700531 | 0 | 0 | 0.12771856 | 0.03067551 |
| 0 | 0.03331357 | 0.06736221 | 0.08049382 | 0.03829323 | 0.07056082 | 0 |
| 0.31270588 | 0.02993136 | 0.04989258 | 0 | 0 | 0.03369079 | 0.04088351 |
| 0.17574026 | 0.07535317 | 0.05562238 | 0.04084582 | 0.00343217 | 0.16274146 | 0 |
| 0.33225748 | 0 | 0.0646199 | 0.02146167 | 0 | 0.03878354 | 0.05676756 |
| 0.19917321 | 0 | 0.0238335 | 0.12167806 | 0 | 0.0361496 | 0.13138357 |

| | | | | | | |
|---|---|---|---|---|---|---|
| 0.19793573 | 0 | 0.04704635 | 0 | 0 | 0 | 0.08535103 |
| 0.12821442 | 0.12695598 | 0.01500505 | 0.00650624 | 0.0149587 | 0.10719447 | 0.04243088 |
| 0.14959561 | 0 | 0 | 0.06363335 | 0.05321093 | 0.04101084 | 0.00615154 |
| 0.32642368 | 0 | 0.0466222 | 0 | 0 | 0.08631679 | 0.03397657 |
| 0.30586239 | 0.06219711 | 0 | 0.02009055 | 0.10658064 | 0 | 0.00198871 |
| 0.15126226 | 0 | 0.0188605 | 0 | 0 | 0.01063346 | 0.06837054 |
| 0.11970835 | 0 | 0.07384409 | 0.00561715 | 0 | 0 | 0.18396416 |
| 0.14962775 | 0 | 0.08373469 | 0.04749834 | 0 | 0 | 0.13827766 |
| 0.23218315 | 0 | 0.03395298 | 0.01760442 | 0.04370663 | 0 | 0.05838899 |
| 0.19850778 | 0 | 0.01649337 | 0.03169228 | 0 | 0.10698218 | 0 |
| 0.24630205 | 0 | 0 | 0.04122057 | 0 | 0.12850692 | 0 |
| 0.06599371 | 0 | 0.03479769 | 0.07636533 | 0.02029856 | 0 | 0.1481911 |
| 0.35226951 | 0 | 0.02832572 | 0 | 0.03898312 | 0.13567971 | 0.07840689 |
| 0.0485189 | 0 | 0.06780209 | 0.12784621 | 0 | 0.05630397 | 0.06582028 |
| 1.06034674 | 0.10397887 | 0 | 0 | 0 | 0.14282448 | 0.24346651 |
| 0.04805995 | 0.00773829 | 0.03368249 | 0.00337203 | 0.00061231 | 0.04863892 | 0 |
| 0.13903289 | 0.0150046 | 0.03431111 | 0.01481248 | 0 | 0 | 0.04942809 |
| 0.10834545 | 0.00336012 | 0.0462073 | 0.00451911 | 0.01214779 | 0.02420061 | 0.0035264 |
| 0 | 0 | 0.06669232 | 0.05956691 | 0 | 0.10282826 | 0 |
| 0 | 0.00398316 | 0.01836651 | 0.00470582 | 0 | 0.11044933 | 0 |
| 0.34233277 | 0.03595532 | 0.02447748 | 0.1035039 | 0 | 0 | 0.03474791 |
| 0.17959652 | 0 | 0.16055212 | 0.22008008 | 0.00683054 | 0.02504947 | 0 |
| 0.1915232 | 0.01374623 | 0.01438152 | 0.03664659 | 0 | 0.07172993 | 0 |
| 0.39005741 | 0 | 0.13346478 | 0.11283713 | 0.06347886 | 0.07428402 | 0 |
| 0.20526826 | 0.00702311 | 0.01526968 | 0 | 0 | 0 | 0.07942021 |
| 0.07547994 | 0 | 0.0365481 | 0.03718362 | 0 | 0.00034008 | 0.09473469 |
| 0.36296248 | 0 | 0.05782925 | 0.0405874 | 0 | 0 | 0.38228172 |
| 0.37723792 | 0 | 0.19637758 | 0.11103338 | 0.00310376 | 0 | 0.61030541 |
| 0.346123 | 0.05945378 | 0.02256232 | 0.0052566 | 0 | 0.01625787 | 0.1018323 |
| 0.154318 | 0 | 0.06486412 | 0.0492245 | 0 | 0.0484389 | 0.0321517 |
| 0.22415408 | 0 | 0.07864591 | 0.1037503 | 0 | 0 | 0.1327168 |
| 0.11908745 | 0 | 0.11488942 | 0.01367496 | 0 | 0 | 0.17933556 |
| 0.33703099 | 0 | 0.08996276 | 0.10096945 | 0 | 0 | 0.17286841 |
| 0.41021463 | 0 | 0.05331068 | 0.19401013 | 0 | 0 | 0.20752528 |
| 0.22524185 | 0 | 0.11421713 | 0.11652595 | 0 | 0.05619474 | 0.12217875 |
| 0.91092748 | 0.05319301 | 0.05869834 | 0.26425288 | 0.00941365 | 0.27003152 | 0 |
| 0.06710113 | 0.00999085 | 0.02347236 | 0 | 0.00333571 | 0.04286615 | 0.02243975 |
| 0.11726024 | 0 | 0.15356353 | 0.03868619 | 0.01571502 | 0 | 0.21006955 |
| 0.38111677 | 0 | 0.04683846 | 0.1488756 | 0 | 0.05607105 | 0.13335637 |
| 0.54929204 | 0 | 0 | 0 | 0 | 0.3025368 | 0 |
| 0.27924854 | 0 | 0.09755287 | 0 | 0 | 0 | 0.13418742 |
| 0.06931863 | 0.01315202 | 0.08304014 | 0.01059621 | 0 | 0.01411867 | 0.03852899 |
| 0.0878173 | 0.05982698 | 0.07140706 | 0 | 0 | 0.14661393 | 0 |

| | | | | | | |
|---|---|---|---|---|---|---|
| 0.08355686 | 0.01635378 | 0.04334829 | 0.01040869 | 0 | 0.06255245 | 0.01338333 |
| 0.1798466 | 0.01156092 | 0.05444379 | 0.01475381 | 0 | 0.01757091 | 0.05171411 |
| 0.06752397 | 0.01085118 | 0.00417609 | 0.00777545 | 0 | 0.05148835 | 0.01973496 |
| 0.13107286 | 0 | 0.03344067 | 0.00933424 | 0 | 0.11567953 | 0.03012374 |
| 0.21951411 | 0 | 0.00803746 | 0.05289031 | 0 | 0.11984558 | 0.00018617 |
| 0.07057202 | 0 | 0.01194821 | 0.00231225 | 0 | 0.01574945 | 0.04746769 |
| 0.28587708 | 0 | 0 | 0.02333025 | 0 | 0.07635348 | 0.09792556 |
| 0.13789509 | 0 | 0 | 0.04231918 | 0 | 0 | 0.09676765 |
| 0.17635263 | 0 | 0 | 0.03090268 | 0 | 0.03462225 | 0.04342144 |
| 0.11935132 | 0.01195182 | 0.00274586 | 0.00540246 | 0 | 0.07850666 | 0 |
| 0.16939489 | 0.00203026 | 0.0243394 | 0 | 0 | 0.05342049 | 0.00160816 |
| 0.09304371 | 0.02379093 | 0.02591544 | 0 | 0 | 0.09138802 | 0 |
| 0.22575475 | 0.05278516 | 0.01832607 | 0.02951135 | 0 | 0.09521443 | 0 |
| 0.47224333 | 0 | 0.05545172 | 0.06229539 | 0.06631992 | 0 | 0.11137855 |
| 0.27686703 | 0 | 0.04953262 | 0.00515883 | 0 | 0.0103835 | 0.08900737 |
| 0.50323393 | 0 | 0.0346464 | 0.02993084 | 0.0124275 | 0 | 0.11757368 |
| 0.36643216 | 0.15378106 | 0.0812135 | 0.14595098 | 0 | 0.23975807 | 0 |
| 0.52068982 | 0.0061698 | 0 | 0.0376489 | 0 | 0 | 0.12262503 |

| Monocytes | Macrophages | Macrophages | Macrophages | Dendritic cell | Dendritic cell | Mast cells re |
|---|---|---|---|---|---|---|
| 0.20338024 | 0.19177231 | 0.02872762 | 0.5592817 | 0.01024267 | 0 | 0 |
| 0.23802775 | 0.48295721 | 0.02792857 | 0.59766453 | 0 | 0 | 0.07899301 |
| 0.1394619 | 0.64151374 | 0.05366169 | 0.4887945 | 0.00188907 | 0 | 0.09070199 |
| 0.32732934 | 0.26565961 | 0.01848806 | 0.39699339 | 0.0245741 | 0.00269511 | 0.06695412 |
| 0.27444606 | 0.19608569 | 0.03453788 | 0.69543995 | 0.01763168 | 0 | 0 |
| 0.1999324 | 0.32572768 | 0.01903494 | 0.8864962 | 0.02753807 | 0 | 0.04245865 |
| 0.53860943 | 0.00457755 | 0.04044725 | 0.5576761 | 0.01314186 | 0.00225072 | 0.02236504 |
| 0.51601828 | 0.32981431 | 0.0139258 | 0.54717173 | 0.02217774 | 0 | 0.08663467 |
| 0.15754936 | 0.81071576 | 0.04749606 | 1.86557841 | 0.0570624 | 0 | 0.29400281 |
| 0.02390354 | 0.94507684 | 0.01998461 | 0.84713113 | 0 | 0.00928616 | 0.10934338 |
| 0.23111669 | 0.41666873 | 0.0147054 | 0.32590835 | 0.01797384 | 0.01816705 | 0.06478981 |
| 0.15795027 | 1.09382981 | 0.07692812 | 1.08627278 | 0 | 0 | 0.12925319 |
| 0.42647991 | 0.06169454 | 0.02043985 | 0.38573097 | 0 | 0.00391642 | 0 |
| 1.28431997 | 0.10872371 | 0.26052725 | 1.19210505 | 0 | 0.06289974 | 0 |
| 0.64963982 | 0.10845342 | 0.08420895 | 0.29773996 | 0 | 0.02007603 | 0 |
| 0.5677792 | 0.26484206 | 0.15338599 | 0.51100541 | 0 | 0 | 0.16828409 |
| 0.89721586 | 0.20109768 | 0.17830535 | 0.66341007 | 0 | 0.01706645 | 0.15193907 |
| 0.42771265 | 0.14447051 | 0.15803666 | 0.54631204 | 0.01639322 | 0 | 0.17206743 |
| 0.59120087 | 0.52179891 | 0.04191828 | 1.59683422 | 0 | 0 | 0 |
| 0.24533236 | 0.73662209 | 0.04333518 | 1.12451146 | 0.01039513 | 0 | 0 |
| 0.37057842 | 0.2941933 | 0.04083928 | 0.73961449 | 0.00708278 | 0 | 0.02523421 |
| 0.37794704 | 0.12638302 | 0.06359299 | 0.48471992 | 0 | 0.00474619 | 0 |
| 0.45412192 | 0.17568514 | 0.03689451 | 0.87849524 | 0 | 0.01661427 | 0.21647234 |
| 0.30781002 | 0.02557759 | 0.04030429 | 0.81405911 | 0 | 2.53E-06 | 0 |
| 0.29327123 | 0.26227744 | 0.05008574 | 0.53267429 | 0.02476095 | 0.00091682 | 0.24741872 |
| 0.40465049 | 0.36720055 | 0.02914826 | 0.63391485 | 0.02173582 | 0 | 0.2526377 |
| 0.62156695 | 0.10792941 | 0.05946856 | 0.7801784 | 0.00128514 | 0.02489595 | 0.22611979 |
| 0.17687598 | 0.73333042 | 0.07092225 | 1.38690595 | 0.03334973 | 0 | 0.29358872 |
| 0.38479432 | 0.20156153 | 0.02544834 | 0.592145 | 0.01736051 | 0.00200737 | 0 |
| 0.41543962 | 0.09976096 | 0.03316093 | 1.41041888 | 0 | 0.07312621 | 0.12905571 |
| 0.52474666 | 0.0678314 | 0.05259717 | 0.93003857 | 0.00029415 | 0.06885905 | 0.29106457 |
| 0.20722993 | 0.5176925 | 0.0727405 | 0.75903103 | 0 | 0 | 0.12501831 |
| 0.44963166 | 0.17211027 | 0.09168962 | 0.49340994 | 0.02820788 | 0 | 0 |
| 1.31774267 | 0.14958643 | 0.04717428 | 0.16091844 | 0 | 0.006439 | 0 |
| 0.41453209 | 0.69813402 | 0.16363798 | 0.88906664 | 0 | 0 | 0 |
| 0.46250122 | 0.18268738 | 0.13966391 | 0.67433243 | 0.00183877 | 0 | 0.14404582 |
| 0.81732363 | 0.30748287 | 0.04870118 | 1.12708156 | 0 | 0 | 0 |
| 0.6621774 | 0.01785106 | 0.04869075 | 1.78373651 | 0.00017814 | 0 | 0 |
| 0 | 1.87572389 | 0.05121494 | 1.34886273 | 0 | 0 | 0 |
| 0.21433077 | 0 | 0.05057812 | 0.35669054 | 0.02143262 | 0 | 0 |
| 0.2049407 | 0 | 0.01931751 | 0.38034854 | 0.01519699 | 0 | 0.08756835 |
| 0.46324009 | 0.00520649 | 0.00749013 | 0.47611713 | 0.02168109 | 0.02457173 | 0.48455673 |

| | | | | | | |
|---|---|---|---|---|---|---|
| 0.43363426 | 0.35986284 | 0.0614268 | 1.52698997 | 0.00341738 | 0 | 0 |
| 0.52795483 | 0.39311272 | 0.13353132 | 1.69248824 | 0.00531149 | 0 | 0.06648949 |
| 0.35302629 | 0.5912557 | 0.04892461 | 1.56740682 | 0.03667069 | 0 | 0.14183095 |
| 0.38828813 | 0.28545919 | 0.06374399 | 1.39421357 | 0.00224421 | 0 | 0.0761458 |
| 0.34353506 | 0.26156877 | 0.23880864 | 0.94106043 | 0.02548734 | 0 | 0.23250474 |
| 0.31480189 | 0.6333669 | 0.06373054 | 1.51848881 | 0.00465374 | 0.02702828 | 0.07355151 |
| 0.25398216 | 0.63816211 | 0.03144848 | 0.65722271 | 0.03861757 | 0 | 0.0815354 |
| 0.71001932 | 0 | 0.01980652 | 0.84451423 | 0.02212684 | 0.00963485 | 0.07736757 |
| 0.56348937 | 0.38735922 | 0.11971953 | 0.79172712 | 0 | 0 | 0.07788569 |
| 0.64162157 | 0.20867052 | 0.03929836 | 0.89056075 | 0 | 0.01275578 | 0 |
| 0.2388954 | 0.40486507 | 0.06526131 | 0.7583294 | 0.00578552 | 0 | 0.4250484 |
| 1.19178185 | 0 | 0.08965415 | 1.50061507 | 0 | 0 | 0.07922226 |
| 1.31282738 | 1.07023205 | 0.01362324 | 0.41276673 | 0.11234804 | 0 | 0.06106943 |
| 0.04863125 | 1.18872415 | 0.0379119 | 0.57121346 | 0 | 0 | 0 |
| 0.75910066 | 0 | 0.20108181 | 1.91344929 | 0 | 0 | 0 |
| 0.56825928 | 0.88835496 | 0.00970343 | 0.84110339 | 0 | 0.02079115 | 0.122886 |
| 0.29867778 | 0.4185538 | 0.0751247 | 0.74897952 | 0 | 0 | 0 |
| 0.30731538 | 0.82528512 | 0.04943326 | 1.45837959 | 0 | 0 | 0 |
| 0.93154702 | 0.03506706 | 0.03011519 | 1.12011686 | 0 | 0 | 0.19443092 |
| 0.19622224 | 0.06752181 | 0.02576118 | 0.65113777 | 0.00145041 | 0 | 0 |
| 0.47603748 | 0.53197296 | 0.04873142 | 1.01652888 | 0 | 0 | 0.00083098 |
| 0.65250095 | 0 | 0.36376888 | 1.69927079 | 0.05794479 | 0 | 0 |
| 0.22863471 | 0.76913315 | 0.02188119 | 0.84899868 | 0.02157218 | 0 | 0.20199402 |
| 0.04930442 | 0.37943684 | 0.03607829 | 0.6101905 | 0.02416256 | 0.00194609 | 0.07142664 |
| 0.43581285 | 0 | 0.06500693 | 0.45150912 | 0.02733628 | 0 | 0.10435605 |
| 0.63337319 | 0 | 0.00963676 | 0.46825507 | 0 | 0.03412169 | 0 |
| 0.058514 | 0.71507976 | 0.0831771 | 1.17176622 | 0.01332841 | 0 | 0.17606793 |
| 1.18118473 | 0 | 0.18561743 | 1.39065641 | 0.03245168 | 0 | 0 |
| 0.57092689 | 0.51454115 | 0.02339547 | 0.70674208 | 0.02150158 | 0 | 0 |
| 0.5911494 | 1.16714288 | 0.07104409 | 1.40556971 | 0 | 0 | 0.10204436 |
| 0.28781518 | 0.05174261 | 0.03465948 | 0.2357187 | 0 | 0.01263096 | 0.08071329 |
| 0.24741085 | 0.15802474 | 0.08817955 | 0.90283581 | 0.01736693 | 0 | 0 |
| 0.04705569 | 0.6926898 | 0.18200965 | 0.65840076 | 0 | 0 | 0.07803108 |
| 0.23894447 | 0 | 0.1099827 | 0.56220161 | 0.03242491 | 0 | 0 |
| 0.80920994 | 0.40815965 | 0.0386788 | 0.83964054 | 0 | 0 | 0.25592196 |
| 0.62337838 | 0.24191051 | 0.121879 | 1.26234904 | 0 | 0 | 0 |
| 0.31058877 | 0.30842703 | 0.05361942 | 0.53567577 | 0.00682336 | 0 | 0 |
| 0.54280542 | 0.11490992 | 0.11899942 | 1.59003702 | 0 | 0 | 0.03834035 |
| 0.05483754 | 0.89147815 | 0.09981427 | 0.9457851 | 0 | 0 | 0.08851091 |
| 0.18975492 | 0.04096421 | 0.0767582 | 0.74514631 | 0.00355782 | 0 | 0.06524233 |
| 0.09362645 | 1.22738379 | 0.02329279 | 0.52237267 | 0 | 0 | 0 |
| 0.12479553 | 0.71537589 | 0.03315083 | 1.02338198 | 0 | 0 | 0.04231592 |
| 0.55991463 | 0.40973927 | 0.03539521 | 0.76201922 | 0.0156288 | 0 | 0.12965275 |

| | | | | | | |
|---|---|---|---|---|---|---|
| 0.2900868 | 0.19876806 | 0.04849532 | 0.64197812 | 0.00117087 | 0 | 0 |
| 0.2646092 | 0.69566739 | 0.12303634 | 2.06510651 | 0 | 0 | 0.08558056 |
| 0.18568774 | 0.18063432 | 0.13298862 | 0.39677274 | 0.03365391 | 0 | 0.07488146 |
| 0.39293914 | 0.43252498 | 0.01719 | 0.94689083 | 0.01271025 | 0 | 0 |
| 0.62480718 | 0.78809903 | 0.02229138 | 1.38173361 | 0 | 0 | 0 |
| 0.33890379 | 0.14893268 | 0.06610098 | 0.99289671 | 0.00337648 | 0 | 0.29480641 |
| 0.53101528 | 0.27933169 | 0.07091341 | 1.20385729 | 0 | 0 | 0 |
| 0.27395243 | 0.09235759 | 0.02553199 | 0.3271453 | 0.00838276 | 0.00760743 | 0 |
| 0.69993792 | 0.29341176 | 0.08845736 | 1.19318452 | 0 | 0 | 0.04523126 |
| 0.22589404 | 0.87342133 | 0.09721475 | 0.60941478 | 0 | 0 | 0 |
| 0.43971027 | 0.4801859 | 0.01694675 | 0.98401829 | 0.02493701 | 0 | 0.08569085 |
| 0.22213409 | 0.70313156 | 0.03341988 | 0.78278196 | 0 | 0 | 0.00958341 |
| 0.32586083 | 0 | 0.01072665 | 0.27342621 | 0.02016496 | 0.02070763 | 0.05507711 |
| 0.37247943 | 0.98590926 | 0.05968227 | 1.01955566 | 0 | 0 | 0.1926545 |
| 1.29807244 | 0.51676574 | 0.00078046 | 0.80119262 | 0.01474923 | 0.11927444 | 0.05704728 |
| 0.12726828 | 0.76711245 | 0.02886405 | 0.8771067 | 0 | 0 | 0.0316773 |
| 0.64143248 | 0.14090902 | 0.0877824 | 1.05822237 | 0.0061409 | 0 | 0.06701443 |
| 0.29195179 | 0.67220583 | 0.08259692 | 0.75406064 | 0 | 0 | 0.06752374 |
| 0.16749398 | 0.81285534 | 0.05337655 | 1.15152602 | 0 | 0 | 0.15496382 |
| 0.05476517 | 0.56455351 | 0.0320036 | 0.91715719 | 0 | 0 | 0.04976073 |
| 0.26315464 | 0.67350608 | 0.10721077 | 1.66630338 | 0 | 0 | 0.14004468 |
| 0.64036101 | 0 | 0.09392324 | 1.67375615 | 0.03377942 | 0 | 0.05823325 |
| 0.48355911 | 0.43192009 | 0.09561859 | 1.58395901 | 0.01038977 | 0 | 0.11770654 |
| 1.15448947 | 2.08162388 | 0.05061186 | 1.26766725 | 0 | 0 | 0.07605204 |
| 0.48736256 | 0.15834886 | 0.06052108 | 0.6782871 | 0 | 0 | 0.12852755 |
| 0.27225363 | 0.12647378 | 0.03869148 | 0.47378041 | 0.00818081 | 0 | 0.12974199 |
| 0.63767656 | 0.0248996 | 0.09527354 | 0.76518871 | 0 | 0 | 0 |
| 1.03058653 | 0.08326978 | 0.22943857 | 1.30343979 | 0 | 0 | 0 |
| 0.58480052 | 0.02441229 | 0.10649785 | 1.19762595 | 0 | 0.01488969 | 0.21456912 |
| 0.28928098 | 0.24228389 | 0.03701556 | 0.61829604 | 0.01445419 | 0 | 0.12513696 |
| 0.41986303 | 0.19508898 | 0.01270542 | 0.50921467 | 0.00734166 | 0.01709004 | 0.02729492 |
| 0.47552111 | 0.06422447 | 0.00817757 | 0.56964767 | 0 | 0.00537724 | 0 |
| 0.28841423 | 0.09758395 | 0.02539665 | 0.70456295 | 7.23E-05 | 0 | 0 |
| 0.41036648 | 0.21857329 | 0.01991623 | 0.33385111 | 0.0148128 | 0 | 0 |
| 0.53116147 | 0.20922649 | 0.01939513 | 0.37626748 | 0.01118802 | 0.01696285 | 0 |
| 0.44979557 | 1.55314476 | 0.44835783 | 3.52705886 | 0.06787173 | 0 | 0.56768081 |
| 0.15041205 | 0.35808696 | 0.05260028 | 0.84529286 | 0.00621737 | 0 | 0.12781672 |
| 0.76704972 | 0.27125687 | 0.14984285 | 0.72341592 | 0 | 0 | 0 |
| 0.65045273 | 0.22707049 | 0.05549148 | 1.09918154 | 0.01840027 | 0 | 0.13156814 |
| 0.19433193 | 2.79819055 | 0.02408446 | 1.81859737 | 0 | 0.01805369 | 0 |
| 0.98688412 | 0 | 0.02424192 | 0.55674802 | 0.01056706 | 0.00872853 | 0 |
| 0.15893419 | 0.4906158 | 0.03758089 | 0.52593298 | 0.01004316 | 0 | 0.05793277 |
| 0.20427547 | 0.44263187 | 0.04358056 | 0.45767895 | 0.01993171 | 0 | 0.01093205 |

| | | | | | | |
|---|---|---|---|---|---|---|
| 0.2151649 | 0.23796693 | 0.04382327 | 0.5056453 | 0.02126198 | 0 | 0.05044665 |
| 0.07538307 | 0.48062065 | 0.03637042 | 0.59871565 | 0 | 0 | 0.04041267 |
| 0.19275283 | 0.09365795 | 0.01804181 | 0.3838261 | 0.0066374 | 0.01207478 | 0.08708772 |
| 0.3375945 | 0.10839238 | 0.07441083 | 0.45980224 | 6.96E-05 | 0 | 0.11037586 |
| 0.28489193 | 0.53895417 | 0.08206726 | 0.84435033 | 0 | 0 | 0.11050691 |
| 0.64994648 | 0.0134728 | 0.01488771 | 0.73724566 | 0 | 0 | 0 |
| 0.89145254 | 0.09913381 | 0.04001487 | 1.21021543 | 0.02334454 | 0.01540174 | 0.19353331 |
| 0.72811561 | 0.01399902 | 0.02106014 | 0.84251632 | 0.01341434 | 0.00911196 | 0 |
| 0.61842009 | 0 | 0.05051251 | 0.50019864 | 0.02335852 | 0 | 0 |
| 0.43572747 | 0.0611563 | 0.01491154 | 0.51317266 | 0 | 0.02059247 | 0.08310846 |
| 0.42501322 | 0.24176311 | 0.05361802 | 0.48271792 | 0 | 0.00674804 | 0.12010621 |
| 0.29541422 | 0.11501351 | 0.02517329 | 0.58306017 | 0 | 0.01180889 | 0.11111728 |
| 0.23360482 | 1.16120394 | 0.03880815 | 0.88443684 | 0 | 0 | 0.14324458 |
| 1.01844702 | 0.19208412 | 0.06172976 | 1.14810582 | 0 | 0 | 0 |
| 0.92226715 | 0.1957853 | 0.05737403 | 0.92171537 | 0 | 0.09711097 | 0 |
| 0.74148298 | 0.0977377 | 0.17710564 | 0.89273773 | 0 | 0 | 0.21558273 |
| 1.46043077 | 0.21298401 | 0.14716254 | 1.38655173 | 0 | 0 | 0.12813726 |
| 0.55578985 | 0.27689506 | 0.24169357 | 1.4043902 | 0 | 0.00153899 | 0.10478504 |

| Mast cells ac | Eosinophils | Neutrophils | T Cells (total) |
|---|---|---|---|
| 0.18440197 | 0.04985534 | 0.01855654 | 0.38757452 |
| 0 | 0.01986968 | 0.02397197 | 0.19336714 |
| 0.01026429 | 0.01276118 | 0.00671162 | 0.18994215 |
| 0 | 0.1222951 | 0.01046339 | 0.25096021 |
| 0.21988418 | 0.0233114 | 0.01569558 | 0.25378775 |
| 0.06875773 | 0.01184553 | 0.03385546 | 0.30748166 |
| 0.06015954 | 0.01870358 | 0.01781912 | 0.33469605 |
| 0 | 0.10128506 | 0.02194059 | 0.24204044 |
| 0 | 0 | 0.01226908 | 1.05354754 |
| 0 | 0 | 0.02572584 | 0.28250835 |
| 0.00419517 | 0.12869012 | 0.08014785 | 0.38257355 |
| 0 | 0 | 0.06682869 | 0.23763885 |
| 0.24229005 | 0.06778897 | 0.00608032 | 0.20703555 |
| 0.56650473 | 0.08240101 | 0.13055583 | 1.34715123 |
| 0.28647329 | 0.12660263 | 0.01424123 | 0.2716493 |
| 0 | 0.1195996 | 0.10570708 | 0.75656608 |
| 0 | 0.019234 | 0.06271624 | 0.56758146 |
| 0 | 0.02899698 | 0.04739244 | 0.5221841 |
| 1.29481232 | 0 | 0.06361041 | 0.41676762 |
| 0.24452034 | 0 | 0.13195142 | 0.55005848 |
| 0.03312334 | 0.04696112 | 0.03738111 | 0.36087129 |
| 0.22590878 | 0.03605598 | 0.01611362 | 0.32949756 |
| 0 | 0.10613664 | 0.00222048 | 0.26860625 |
| 0.24641327 | 0.03320409 | 0.04148289 | 0.50556368 |
| 0 | 0.06331493 | 0.00159888 | 0.54047855 |
| 0 | 0 | 0.03128547 | 0.25691452 |
| 0 | 0.10606885 | 0.00878939 | 0.32450458 |
| 0 | 0.00356043 | 0.03565249 | 0.61439873 |
| 0.09002963 | 0.04088146 | 0.05224247 | 0.25451183 |
| 0 | 0 | 0 | 0.73799653 |
| 0 | 0.01691011 | 0.0054309 | 0.6263536 |
| 0 | 0.01831828 | 0.02197992 | 0.50260122 |
| 0.05718162 | 0.06217696 | 0.01756435 | 0.39472116 |
| 0.90629113 | 0.37582862 | 0.67593824 | 1.02006075 |
| 0.49946725 | 0.02149062 | 0.14207504 | 0.56509596 |
| 0 | 0.01706052 | 0.01429993 | 1.0507764 |
| 0.90175005 | 0.02567477 | 0.05076428 | 0.35170884 |
| 0.17861714 | 0 | 0.04360689 | 0.65233353 |
| 0.23420635 | 0.00512561 | 0.10116371 | 1.11770607 |
| 0.80368586 | 0 | 0.03659966 | 0.33411232 |
| 0.3533844 | 0 | 0.05405619 | 0.30173934 |
| 0 | 0 | 0.00788799 | 0.13740373 |

| | | | |
|---|---|---|---|
| 0.03906565 | 0.01628468 | 0 | 0.20606739 |
| 0 | 0.00340483 | 0.00353796 | 0.42671731 |
| 0 | 0.00890565 | 0.04803888 | 0.6364738 |
| 0 | 0.00891905 | 0.00046605 | 0.22865384 |
| 0 | 0.00510909 | 0.04591206 | 0.6858945 |
| 0.02092486 | 0.02045874 | 0.02561583 | 0.62814119 |
| 0.0036789 | 0 | 0.03995551 | 0.50008963 |
| 0 | 0.02419735 | 0.01237394 | 0.24777574 |
| 0 | 0.26702018 | 0.02853715 | 0.26139825 |
| 0.08121329 | 0 | 0.08968191 | 0.62240059 |
| 0 | 0.05087053 | 0.00104451 | 0.30123086 |
| 0 | 0 | 0.05577334 | 0.32253561 |
| 0 | 0.01300305 | 0 | 0.56797282 |
| 0.07328344 | 0.00957712 | 0.07553337 | 0.22249217 |
| 0.20002677 | 0 | 0.01189068 | 1.41742144 |
| 0 | 0 | 0.16476575 | 0.23891542 |
| 0.07514384 | 0 | 0.04334406 | 0.87504684 |
| 0.54408193 | 0.16757003 | 0.02485008 | 0.36096208 |
| 0 | 0 | 0.00035246 | 0.56612007 |
| 0.07134557 | 0.01193636 | 0.06281418 | 0.25915835 |
| 0.04493091 | 0 | 0.04505805 | 0.44959542 |
| 0.12357655 | 0 | 0.02593573 | 2.03823119 |
| 0 | 0.00400719 | 0.0696713 | 0.32475183 |
| 0 | 0.15444669 | 0.17609329 | 0.82473493 |
| 0.04906517 | 0.04651662 | 0.01603642 | 0.27936119 |
| 0.17451689 | 0.09304568 | 0.11112711 | 0.52124525 |
| 0 | 0 | 0.08355079 | 0.61528761 |
| 1.30447604 | 0 | 0.03136247 | 1.40660699 |
| 0.06120075 | 0.04183861 | 0.00160782 | 0.57043979 |
| 0 | 0 | 0.07628124 | 0.86748733 |
| 0 | 0.09209972 | 0 | 0.23074923 |
| 0.23574278 | 0.01111282 | 0.01634086 | 0.32452953 |
| 0 | 0.02326835 | 0.01488575 | 0.3675453 |
| 0.06912475 | 0.03416846 | 0.01948397 | 0.33076683 |
| 0 | 0.05735994 | 0.06364174 | 0.48257843 |
| 0.05354017 | 0.06418893 | 0.13476909 | 0.477042 |
| 0.20422545 | 0.07572645 | 0.02782498 | 0.26842164 |
| 0.00989658 | 0 | 0.07384345 | 0.57577507 |
| 0 | 0.0052078 | 0.14028787 | 0.28897953 |
| 0 | 0.04964115 | 0.00060823 | 0.39252982 |
| 0.0907556 | 0.03150217 | 0.0211101 | 0.3509938 |
| 0 | 0.03974874 | 0.02228958 | 0.44663829 |
| 0 | 0 | 0.00154592 | 0.71841495 |

| | | | |
|---|---|---|---|
| 0.18439774 | 0.07888422 | 0.02831364 | 0.44434917 |
| 0 | 0.0021966 | 0.01288221 | 0.45496025 |
| 0 | 0.03747406 | 0.04499818 | 0.27896909 |
| 0.54924876 | 0.08999012 | 0.05854434 | 0.42407048 |
| 0.07738564 | 0.01551751 | 0.0199816 | 0.55072015 |
| 0 | 0.04223242 | 0.08685387 | 0.32290365 |
| 0.07784994 | 0.0272933 | 0.05458064 | 0.47533603 |
| 0.36347902 | 0.03258494 | 0.0090961 | 0.39665563 |
| 0 | 0.05145192 | 0.04992354 | 0.51091843 |
| 0.05532581 | 0.02061922 | 0.03514787 | 0.24975721 |
| 0 | 0.13113801 | 0.13932934 | 0.36428416 |
| 0.05316995 | 0.00447522 | 0.01188545 | 0.26997309 |
| 0.00410442 | 0.03739726 | 0.03929786 | 0.53397733 |
| 0.0016685 | 0 | 0.04349985 | 0.369378 |
| 0 | 0 | 0 | 1.27091684 |
| 0.02614546 | 0.03420369 | 0.02188023 | 0.16964407 |
| 0 | 0.04756743 | 0.00410488 | 0.32779575 |
| 0 | 0.01208132 | 0.01565003 | 0.2153326 |
| 0 | 0.01245684 | 0.01638169 | 0.24486958 |
| 0.00263888 | 0 | 0.21099173 | 0.19825582 |
| 0 | 0 | 0.03465856 | 1.02459342 |
| 0 | 0.09344464 | 0.02348658 | 1.26265334 |
| 0 | 0.01763124 | 0.02444176 | 0.52880038 |
| 0.80082368 | 0 | 0.15659015 | 0.77443884 |
| 0 | 0.05721289 | 0.00119594 | 0.25491029 |
| 0 | 0.03426934 | 0.01448525 | 0.26225285 |
| 0.12876741 | 0.02114069 | 0.0191253 | 0.53566437 |
| 0.87323603 | 0.02752732 | 0 | 0.84637376 |
| 0 | 0 | 0 | 0.46563433 |
| 0 | 0.01544049 | 0.0174444 | 0.32352576 |
| 0.04447213 | 0.00588725 | 0.10859799 | 0.56065688 |
| 0.28602629 | 0.00829814 | 0.20817699 | 0.31750181 |
| 0.18404959 | 0.0016595 | 0.08679693 | 0.64876434 |
| 0.06250989 | 0.00890776 | 0.17646469 | 0.75557077 |
| 0.21329226 | 0.01897812 | 0.08569657 | 0.63640703 |
| 0 | 0 | 0.08965101 | 1.89939872 |
| 0 | 0.01772272 | 0.0354114 | 0.14508165 |
| 0.3960381 | 0.03941549 | 0 | 0.50671345 |
| 0 | 0.01922571 | 0.04205098 | 0.73585209 |
| 0.20524851 | 0 | 0.11949888 | 0.75791815 |
| 0.86265925 | 0.04448451 | 0.04129417 | 0.51918276 |
| 0.00169563 | 0.0354641 | 0.01302484 | 0.22594564 |
| 0.06598951 | 0.03691219 | 0.01728217 | 0.29255831 |

| | | | |
|---|---|---|---|
| 0.01880898 | 0.0364684 | 0.01823291 | 0.22643046 |
| 0.02054087 | 0.03119138 | 0.04893771 | 0.33884568 |
| 0 | 0.00897166 | 0.00122925 | 0.16189357 |
| 0 | 0.04191051 | 0 | 0.25582214 |
| 0 | 0.05433109 | 0.0519842 | 0.35068528 |
| 0.10524519 | 0.01444306 | 0 | 0.12097433 |
| 0 | 0.02795953 | 0 | 0.3271571 |
| 0.15020225 | 0.02440898 | 0.01865044 | 0.33725125 |
| 0.26093723 | 0.02676406 | 0.05644731 | 0.31123939 |
| 0 | 0.02329036 | 0.01215065 | 0.1830938 |
| 0 | 0.00981035 | 0.01400999 | 0.31472844 |
| 0 | 0.02257872 | 0.00464028 | 0.23989286 |
| 0 | 0.0053082 | 0.06959963 | 0.37189533 |
| 0.17702822 | 0.0125622 | 0.0996368 | 1.02823127 |
| 0.34252941 | 0.02527712 | 0.10177721 | 0.37667463 |
| 0 | 0.008599 | 0.03154326 | 0.77329262 |
| 0.00740644 | 0 | 0.13542821 | 1.28028982 |
| 0 | 0 | 0.20649872 | 0.70299317 |
